# Integrative multi-omics reveal glial signatures associated with accelerated cognitive decline in Alzheimer’s disease

**DOI:** 10.1101/2024.08.27.24312641

**Authors:** Eléonore Schneegans, Nurun Fancy, Michael Thomas, Emily Adair, Nanet Willumsen, Marianna Papageorgopoulou, Vicky Chau, To Ka Dorcas Cheung, Robert C.J. Muirhead, Harry Whitwell, Riad Yagoubi, Xiaowen Zhang, Aisling McGarry, Brian M Schilder, Paul M. Matthews, Johanna S Jackson

## Abstract

Alzheimer’s disease (AD) is a neurodegenerative disorder characterised by progressive cognitive decline and memory loss caused by both genetic and environmental factors. Synapse dysfunction and loss are strongly related to cognitive decline in AD. This study integrates genomic, transcriptomic, proteomic and immunohistological (multi-omics) data and longitudinal cognitive data across several AD cohorts to elucidate the molecular drivers associated with astrocytes and microglia involved in these processes. Our findings demonstrate that activation of microglia and astrocytes occurs in specific cell subsets that are enriched in AD risk genes. Transcriptomic evidence for early microglial activation precedes immunohistological expression of severe neuropathology. Proteomic markers of astrocytic response appear to be most strongly associated with accelerated cognitive decline. However, we also found that brains from donors with a history of more rapid cognitive decline showed evidence for reduced SNAP25-VAMP interactions indicative of synaptic dysfunction, exhibited higher neurotoxic astrocyte reactivity, and were associated with the expression of neuronal markers of injury. Related molecular signatures in cerebrospinal fluid and plasma may provide biomarkers to identify patients at higher risk for rapid cognitive decline. Together, our results connect glial activation to synaptic dysfunction and cognitive decline in AD and highlight roles for microglial activation in the genesis of AD and later astrocyte activation as a potential determinant of clinical symptom progression.

## Introduction

The neuropathological hallmarks of Alzheimer’s Disease (AD), amyloid-beta (Aβ) plaques and neurofibrillary tau tangles (NFT), are associated with a range of molecular cell pathologies and neurodegeneration. However, cognitive decline remains the most clinically relevant endpoint; it is directly and proximately related to patient quality of life and perceived disease burden^1,2^. This decline, alongside neuropathology, necessitates a deeper exploration of AD beyond amyloid and tau, prompting the need for integrating cognitive data with high-throughput omics techniques to elucidate the cellular and molecular events preceding and accompanying cognitive symptoms. *Post mortem* brain data resources that include multiple types of omics data from ROSMAP^3^, MSBB^4^, and the UK DRI Multi-omics Atlas Project (MAP) are facilitating the elucidation of AD molecular pathology^5,6^. ROSMAP and MSBB cohorts, which are also clinically well-phenotyped longitudinally during the donors’ lifetimes, provide cognitive performance data that can be linked to the *post mortem* ‘omics measures.

The cellular phase of AD, characterised by glial activation and vascular changes before noticeable cognitive deficits emerge, is now at the forefront of research. This focus is due to the promise afforded by understanding and then therapeutically targeting early mechanisms likely to be causal in the disease^7^. Single-nuclei RNA-sequencing (snRNA-seq) has significantly advanced our understanding of AD pathogenesis by profiling diverse microglial^8,9^ and astrocyte^10–13^ phenotypes associated with AD progression. These have defined glial states exhibiting both neuroprotective and neurotoxic features^9,13^. Most AD-associated risk genes identified through Genome-Wide Association Studies (GWAS) converge on microglial response pathways^14^ related to responses to Aβ accumulation. Associated functions played by sub-states have roles in Aβ clearance and brain homeostasis^9^. These have shown, for example, that the TREM2 receptor is pivotal in microglial phagocytosis of Aβ and acts as an immune modulator. A loss-of-function variant (*TREM2*^R47H^) was associated with early neuronal dysfunction^12^ and reduced synaptic density with the upregulation of several complement cascade components^17^. Impaired amyloid clearance has also been associated with activation of astrocytes^15^. Normally playing central roles in synapse formation and maintenance^16^, with AD pathology they undergo reactive astrogliosis in response to AD pathology, characterised by morphological changes and increased expression of immune response genes^17,18^. These reactive astrocytes can increase Aβ production, impair its clearance^19^, and release proinflammatory molecules with neurotoxic oxidative stress, synapse elimination and neurodegeneration^19^. These latter processes appear to be associated proximately with cognitive decline^20^.

Defining the pseudo-temporal evolution of glial activation and related cellular changes in neurons can provide a pathological foundation for the progression of the clinical disease ^21,22^. Here, we have investigated relationships between cognitive decline in AD with molecular signatures of microglia and astrocytes defined from an integrative multi-omics approach, including AD polygenic risk (genomics), untargeted bulk transcriptomics-proteomics, single-cell transcriptomics, targeted presynaptic proteomics, and cerebrospinal fluid (CSF)/plasma proteomics across multiple human AD cohorts (***Supplementary Fig 1***). We employed pseudotemporal analyses to characterise the evolution of molecular pathology and to characterise its associations with disease progression. Our findings indicate that early microglial response is cell subtype-specific, directly modulated by genetic risk and antecedent to significant neuropathology. Subsequently, a secondary response mediated by astrocytic reactivity is associated with a greater rate of clinical progression and neuropathology load but is not directly influenced by AD genetic risk. We provide evidence for specific glial responses with synaptic dysfunction in early-stage AD. The presence of corresponding molecular signatures in the CSF and plasma suggests their potential as biomarkers for patient stratification.

## Results

### Bulk transcriptomics-proteomics integration defines AD-correlated glial activation and neuronal injury responses

We included bulk transcriptomics and proteomics data derived from 112 samples acquired from the middle temporal gyrus (MTG) and somatosensory cortex (SOM) of 56 *post mortem* brains from neuropathologically diagnosed AD and non-diseased control (NDC) donors of comparable ages (n=33 AD & n=23 NDC, mean (SD) 80.4 (10.8) & 80.5 (8.13) years old, ***Supplementary Table 1***). Our analysis involved an unsupervised integration of bulk transcriptomics and proteomics data through the Multi-Omics Factor Analysis (MOFA) model^23^ to identify shared axes of variation (i.e. *factors*) relevant to AD pathology. We found that a model using K=15 factors explained 69% of the variation in the transcriptomics data and 23% in the proteomics data (***Supplementary Table 3, Supplementary Fig 2***). Expression of factors 1,7,11, and 14 were correlated with measures of AD neuropathology, β-amyloid [4G8] and pTau [PHF1] (range of R^2^ value from 0.26 to 0.46, *p≤0.05,* ***Fig 1 a***). We found that factor 3 variance was well explained by brain region (R^2^=0.81, *p≤0.001)*.

**Figure 1.**
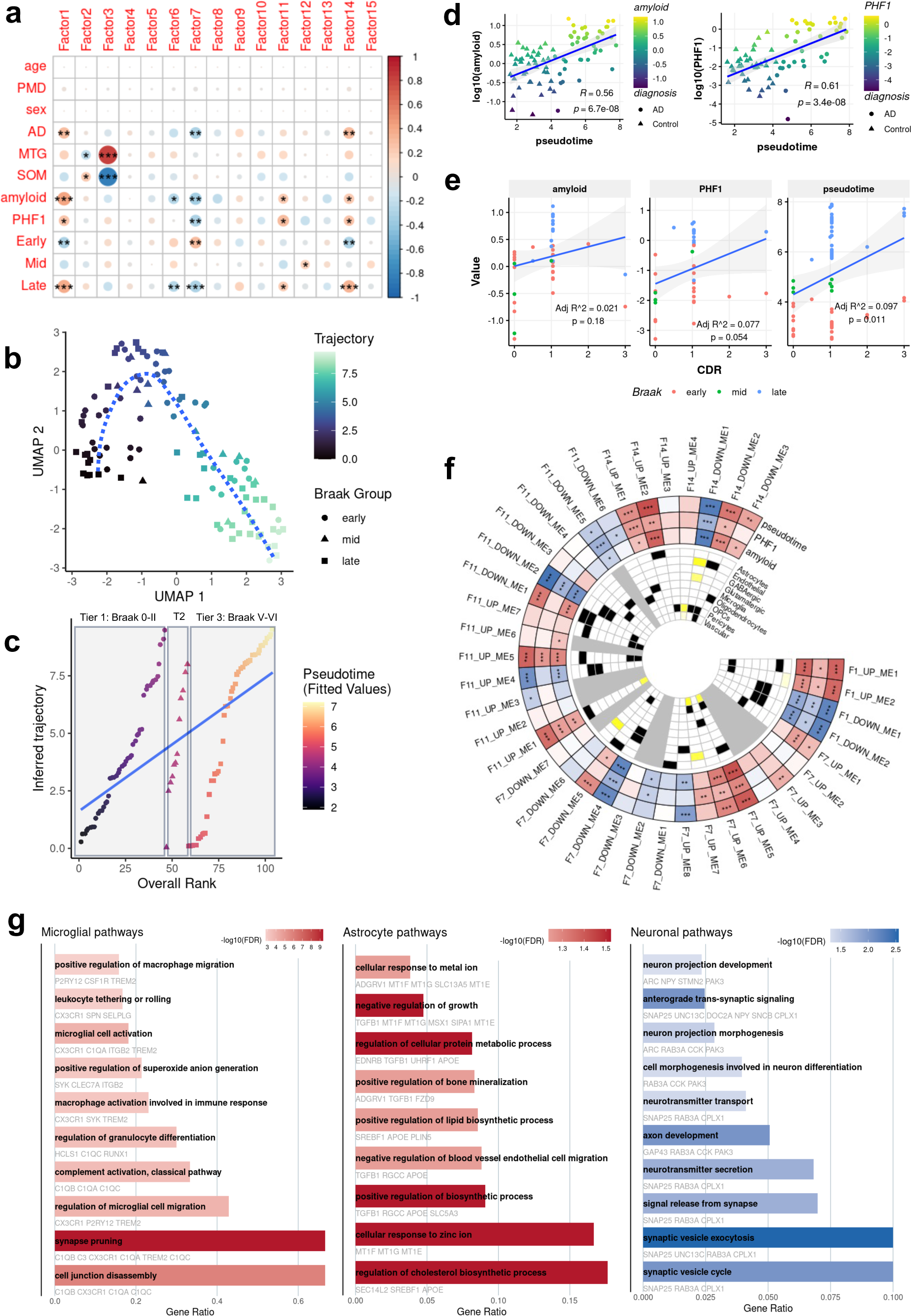
Bulk transcriptomics-proteomics integration defines AD-correlated glial activation mechanisms and neuronal injury response. ***a)*** Heatmap showcasing the Pearson’s correlation matrix of various clinical and pathological parameters with MOFA factors derived from bulk transcriptomics-proteomics integration. Age, PMD and sex have been regressed before integration. ***b)*** Uniform Manifold Approximation and Projection (UMAP) plot illustrating the distribution of brain tissue samples based on a trajectory derived via Slingshot and AD progression. Each point represents a tissue sample coloured by trajectory, ranging from early (black) to late (light cyan) stages of the disease**. c)** This plot illustrates the trajectory of AD progression plotted against the ranked positioning of brain tissue samples according to Braak staging criteria into tiers. The x-axis represents the overall rank of samples from early to late Braak stages, while the y-axis denotes the trajectory inferred through pseudotime analysis. The linear model depicts the correlation between sample ranking within the tiers and disease trajectory. Fitted values correspond to the final pseudotime value. **d)** Linear regression fit of log_10_(amyloid) against pseudotime showing a significant association (left). Linear regression of log_10_(PHF1) against pseudotime showing a significant association (right). **e)** Linear regression fit of pseudotime / log_10_(PHF1) / log_10_(amyloid) against CDR. Points coloured by Braak group: early (0-II), mid (III-IV), late (V-VI) **f)** Circos plot visualising module trait relationships Pearson’s correlations indicating significant associations with key AD features (positive association in red, negative in blue). Cell type enrichment in the inner circle indicates significant enrichment in black (p<0.05), near significance in yellow (0.05<p<0.1), and NS in white. Pearson’s correlation p-values derived from t-test (*p≤0.05, **p≤0.01, ***p≤0.001) **g)** Pathway enrichment analysis (GO Biological Process) for selected up-regulated top microglia (F14_UP_ME1) and astrocyte (F14_UP_ME2) modules and the top downregulated neuronal (synaptic) module (F11_DOWN_ME2), with colour intensity representing the -log_10_(FDR) and key genes highlighted within each pathway.

These AD-correlated factors and the brain region-relevant factor were then used in a semi-supervised approach to derive a brain region-dependent disease pseudotime trajectory (***Fig 1 b,c***). Along this trajectory, pseudotime was correlated with both local tissue phospho-tau (PHF1, R^2^=0.61, P<0.001) or β-amyloid (4G8, R=0.56, P<0.001) immunostaining area (***Fig 1 d)***. We also found that clinical disease progression measured on the Clinical Dementia Rating Scale (CDR)^2^ was associated with the pseudotime trajectory. In models where pseudotime, amyloid, or PHF1 were regressed against CDR, the pseudotime model provided the best explanation of variance (Adj R^2^= 0.097, ***Fig 1 e***).

We constructed transcriptomics-proteomics co-expression networks based on features contributing to the AD-correlated factors and identified co-expression modules that we associated with AD (***Fig 1 f, Supplementary Tables 4-5***). We then identified the 15 most highly differentially expressed modules with greater pseudotime (***Supplementary Fig 3, Supplementary Tables 6-7***). We validated the association of these modules with AD by showing that the eigenvalues of each module were significantly differentially expressed with AD compared to controls in the independent MSBB cohort dataset (***Supplementary Fig 4***).

To determine brain cell types most strongly associated with each module, we used EWCE^24^ to test for their relative enrichment with cell-type data reference from the Allen Human Brain Atlas^25^. Up-regulated modules were enriched for genes most strongly expressed in endothelial cells, oligodendrocytes, astrocytes, and microglia. Down-regulated modules were enriched for genes expressed in GABAergic and glutamatergic neurons (***Fig 1 f***). We selected three modules with the greatest fold change along pseudotime and the most significant associations with microglia, astrocytes, and neurons (***Supplementary Fig 3***): one up-regulated module enriched for microglia gene and protein expression (F14_UP_ME1), one up-regulated module similarly enriched for astrocytes (F14_UP_ME2) and one down-regulated module enriched for neurons (F11_DOWN_ME2). Functional enrichment analysis linked each module to distinct pathology-related pathways. F14_UP_ME1 (microglial) was associated with inflammatory response and synaptic pruning, F14_UP_ME2 (astrocytic) with metal ion homeostasis, and F11_DOWN_ME2 (neuronal) with synaptic functionality and exocytosis (***Fig 1g, Supplementary material Pathway Reports***). We interpret these as mechanistic pathways expressed with AD with neuropathology and clinical progression (***Supplementary Fig 5***).

### Microglia and astrocyte activation with AD is cell state-specific and enriched in AD risk genes

In our previous work^26,27^, we annotated snRNA data into distinct cell types whose samples partly overlap with the current bulk cohort (42 out of 112 samples). Microglia, astrocytes and neurons were kept for the present analysis and further sub-clustered into cell subtypes. Here, we employed high-dimensional Weighted Gene Co-expression Network Analysis (hdWGCNA) to project the previously identified modules onto the sub-clustered microglia, astrocytes and neuronal snRNA data. This allowed cell and cell sub-type specific expression of genes to be identified within the bulk modules. Specifically, within the microglia population, we identified the Micro2 subtype, which here showed cell subtype-specific expression patterns from the projected microglia bulk modules (***Fig 2 a-b***) and DAM and HLA-like states^9^ (***Fig 2 c, Supplementary Fig 6***). Astro0 and Astro2 showed cell subtype-specific expression patterns from the projected astrocyte bulk modules, with Astro0 coinciding with F14_UP_ME2 (enriched for metal ion homeostasis) and Astro2 with F11_UP_ME1 (enriched in glutathione metabolism) (***Fig 2 d-e, Supplementary Files 2-3***). Astro0 showed higher expression of neurotoxic genes, while Astro2 was a neuroprotective gene expression signature (***Fig 2 f, Supplementary Fig 6, Supplementary Table 8***). Neurons showed higher expression in specific neuronal subtypes, notably including RORB^+^ excitatory subpopulations (***Fig 2 g-h, Supplementary Fig 7***). Enrichment analyses using MAGMA^28^ and AD GWAS^29–31^ studies in a meta-analysis were used to assess enrichments for AD genetic risk in microglial and astrocytic bulk modules separately for the transcriptomic or proteomic data alone or for the union of the features. We found that DAM/HLA-associated microglia (F14_UP_ME1/Micro2) signatures were significantly enriched in AD risk genes and that this was most strongly associated with the transcriptomic signatures (***Fig 3 a-b***). The proteomics signature for the neurotoxic astrocyte module (F14_UP_ME2/Astro0) was also enriched for the expression of AD risk genes (***Fig 3 a,c***). Through transcription factor (TF) target gene enrichment analysis, we discovered significant enrichment of STAT1 (OR=3.7, p-value=2e-03) and CEBPB (OR= 4.1, p-value= 2e-04) target genes in the microglia module, underscoring its potential as a key regulatory hub in transcriptional networks. A similar analysis of the astrocyte module did not identify significant enrichment for transcription factors. We hypothesise that the proteomic signature may be regulated predominantly post-transcriptionally (***Fig 3 d***).

**Figure 2.**
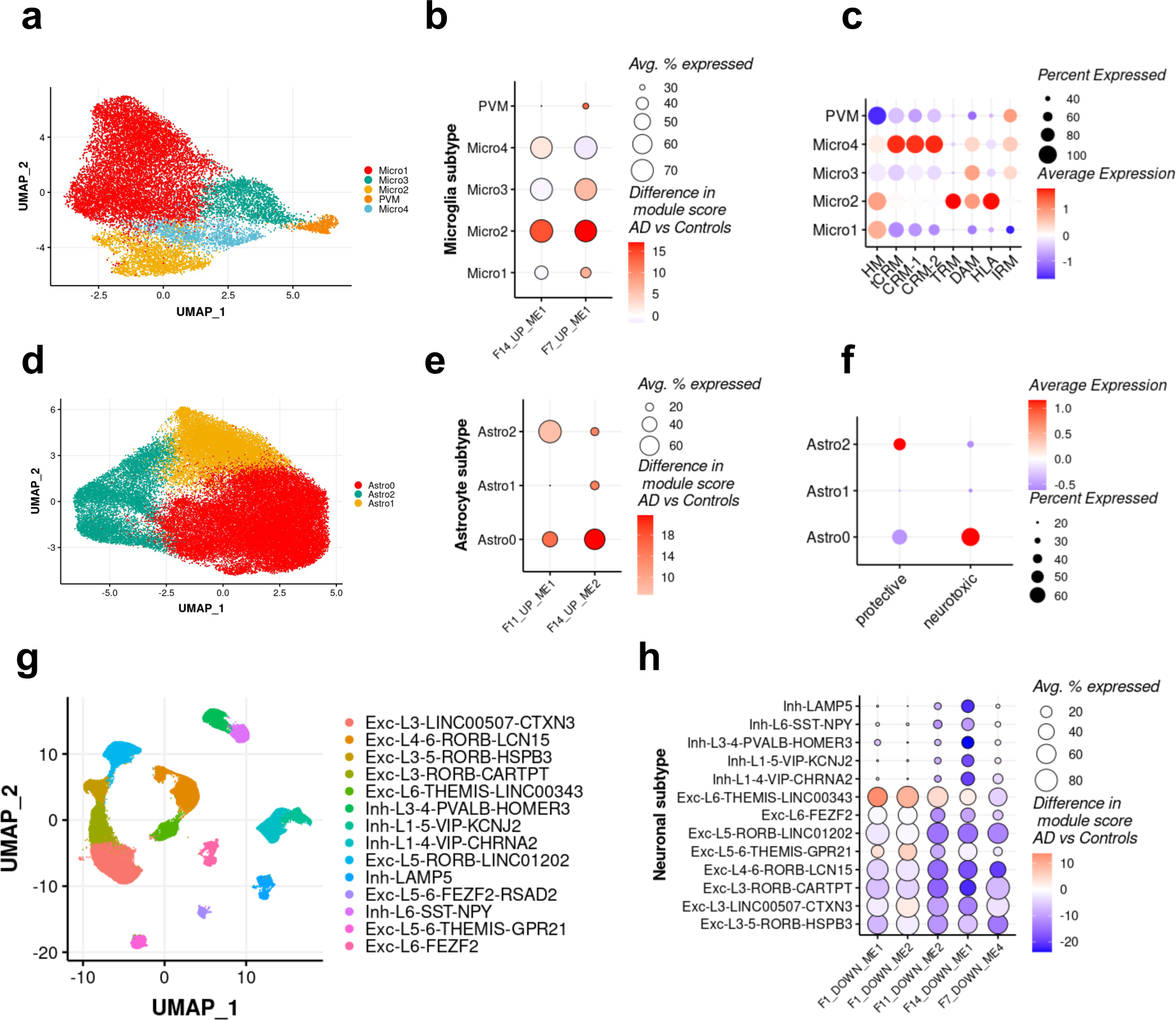
Functional characterisation of bulk modules within their sub-cellular context. **a)** UMAP visualisation of microglial subpopulations in snRNAseq**. b)** Projecting AD-correlated microglia modules on single-cell clusters using hdWGCNA with colour intensity indicating the difference in module score between AD patients and NDC. The micro2 cluster is specific to the two microglial-enriched bulk modules, F14_UP_ME1 and F7_UP_ME1. Correspondence is assigned to the module-subpopulation pair with the most significant average expression and greatest difference in module score in AD vs Controls. **c)** Dot plots quantifying the average percentage expression of Mancuso et al. defined microglial subtypes^9^. Colour gradient representing scaled average expression levels. **d)** UMAP visualisation of astrocyte subpopulations in snRNAseq**. e)** Same as in b but using astrocyte-correlated modules on astrocyte subpopulations. Astro0 and Astro2 correspond to F14_UP_ME2 and F11_UP_ME1 respectively. **f)** Dot plots quantifying the average percentage expression of Cameron et al. astrocyte subtypes^13^. **g)** UMAP visualisation of neuronal subpopulations in snRNAseq**. h)** Same as in b/e but using neuronal-correlated modules on neuronal subpopulations.

**Figure 3.**
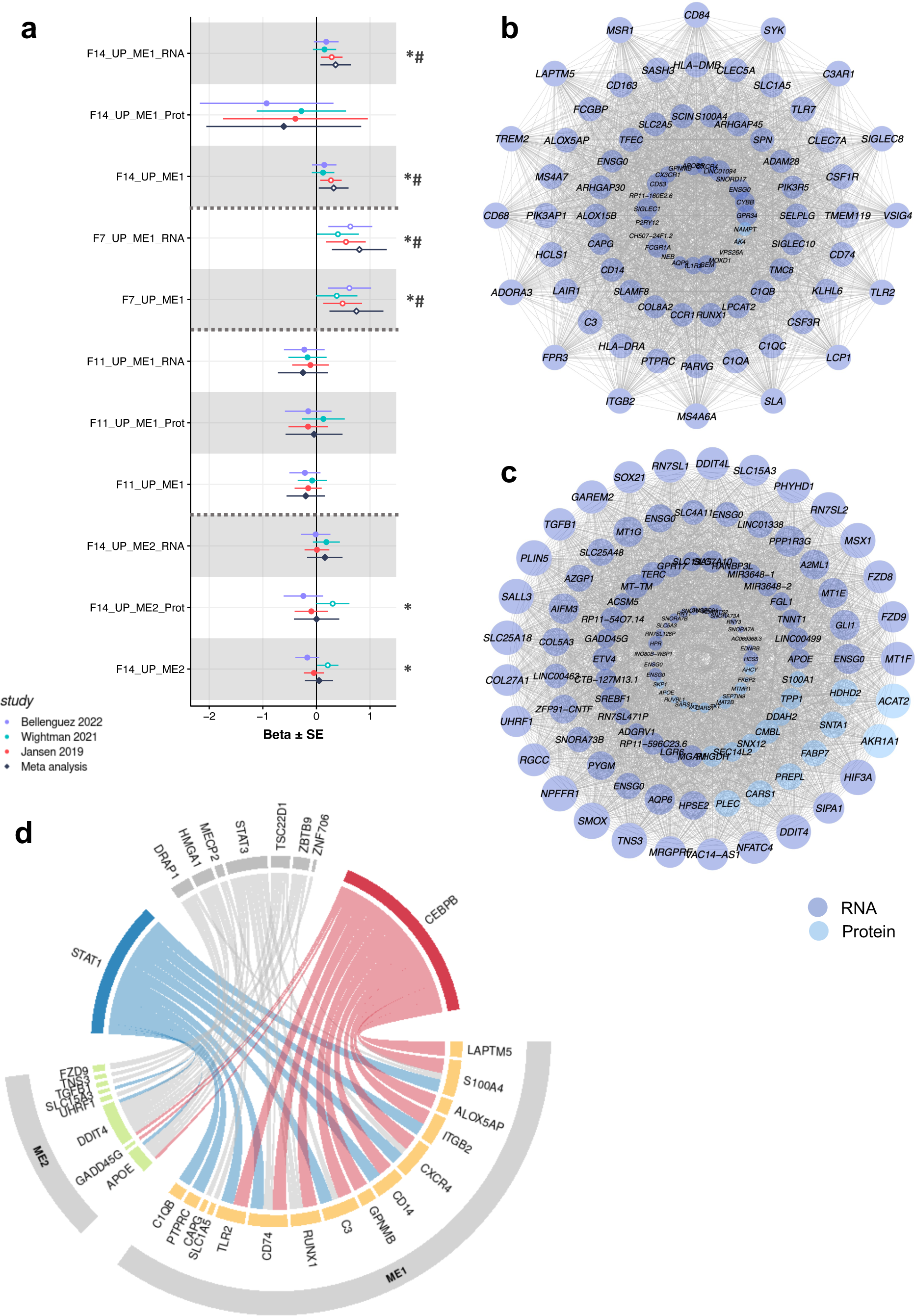
Microglia and astrocyte activation mechanisms are enriched in AD genetic risk. **a)** Forest plot illustrating the enrichment of microglia (F14_UP_ME1, F7_UP_ME1) and astrocyte co-expression modules (F14_UP_ME1, F11_UP_ME1) (split by transcriptomics ‘_RNA’, proteomics ‘_Prot’, and union of features) in genetic risk quantified by three AD GWAS ^29–31^ and in a meta-analysis, highlighting significant beta values and corresponding P-values. # indicates modules enriched in AD risk genes in the meta-analysis and * only in one or more studies. **b-c)** Schematic co-expression network diagrams showing module membership of genes (dark blue) and proteins (light blue) in F14_UP_ME1 (top) and F14_UP_ME2 (bottom). Node size is proportional to connectivity (degree), and nodes are separated into four quantiles representing each layer. **d)** Integrated transcriptomics-proteomics analysis uncovers disease progression-specific TF regulatory mechanisms in microglia and astrocytes. Circos plot displays TF proteins and their target genes in ME1 (F14_UP_ME1) microglia and ME2 (F14_UP_ME2) astrocytic modules. TFs with significant enrichment of target genes in ME1 are shown in blue (TF with negative factor 14 weight) and red (TF with positive factor 14 weight). Other TF with no significant enrichment of target genes in modules are indicated in grey.

### AD polygenic risk modulates glial activation and its effects on the rate of cognitive decline

To explore the genetic influence on glial activation and clinical disease in early AD, we computed polygenic risk scores (PRS) derived from SNPs in genes within microglial and astrocyte activation modules that we had shown to be enriched for AD risk (***Supplementary Fig 8, Supplementary Tables 9-11***). PRS scores were computed for the subset of data from donors in the ROSMAP cohort with early AD (Braak stages III-IV, *n*=132). The microglial PRS (***Fig 4 a***) was significantly correlated with microglial module eigenvalue, consistent with genetic influence on microglial activation (***Fig 4 c***). Despite this genetic influence on microglial module eigenvalue, microglial PRS did not significantly correlate with neuropathological measures or the rate of clinical progression. Conversely, the astrocyte PRS, featuring SNPs from genes including *PLEC* and *APOE* (***Fig 4 b***), did not directly correlate with astrocyte module eigenvalue but was correlated with Aβ burden (expressed as amyloid load, diffuse or neuritic plaques counts) (***Fig 4 c***). Increased astrocyte eigenvalue was significantly associated with neuropathology and a faster rate of cognitive decline (***Fig 4 c***). Moreover, astrocyte and microglia eigenvalues were positively associated with pseudotime. Together, these results suggest that genetic risk is a determinant of microglial activation in AD. By contrast, we hypothesise that astrocytic activation responses with AD, which appear to have a lower genetic association, are related to local environmental signals.

**Figure 4.**
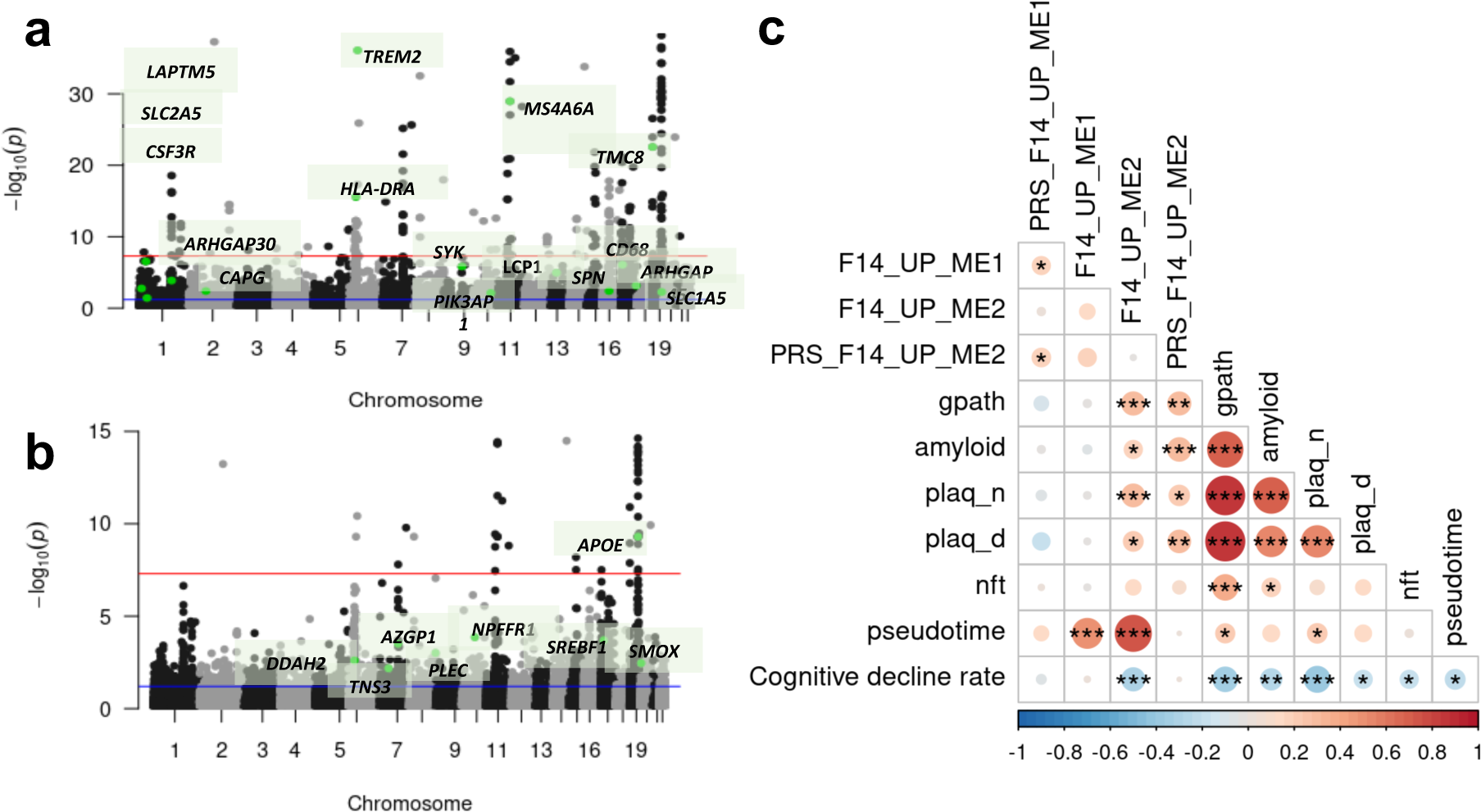
AD polygenic risk modulates glial activation directly and indirectly. **a-b)** Manhattan plots displaying the MAGMA gene-based association test results for AD-correlated modules F14_UP_ME1 (microglia) and F14_UP_ME2 (astrocyte), with genome-wide significance thresholds indicated and notable genes highlighted. The red line indicates genome-wide significance (p≤5x10^-8^) and the blue line (p≤0.05). **c)** Pearson’s correlation heatmap of microglia and astrocyte PRS, modules, and neuropathology measures, including a global pathology measure (quantitative summary of AD pathology derived from counts of three AD pathologies: neuritic plaques (n), diffuse plaques (d), and neurofibrillary tangles (NFT), and rate of cognitive decline in the last three years preceding death. Pearson’s correlation p-values derived from t-test (*p≤0.05, **p≤0.01, ***p≤0.001).

### Glial activation is associated with neuronal signatures of synapse dysfunction

We tested whether the glial activation mechanisms identified in astrocytes and microglia—enriched for AD risk genes—and the neuronal injury response module were associated with accelerated cognitive loss in the early stages of AD (Braak III-IV). Utilising the ROSMAP cohort in early AD, we separated individuals into two subgroups based on unsupervised multi-omics clustering^32^ (***Supplementary Fig 9, Supplementary Tables 12-13***). After the exclusion of individuals exhibiting a survivor bias (those with follow-up years>12, ***Supplementary Fig 9***c), a Kaplan Meier analysis uncovered a group with faster progression to AD (blue) and a group with a slower clinical progression (yellow), with no significant demographic or AD pathology differences between them (***Fig 5 a***, ***Supplementary Table 13***). We also analysed the global cognitive function, which is derived from the average of 19 cognitive tests. These tests span multiple domains, including episodic, semantic and working memory, visuospatial ability and perceptual speed. Most ROSMAP participants have taken these tests annually, allowing for a detailed longitudinal assessment of cognitive changes over time. When examining longitudinal cognitive profiles, we observed significant differences in the rate of cognitive decline between these groups, where the fast group showed an accelerated rate of decline compared to the slow group (***Fig 5 b***). Further downstream analysis uncovered that it was primarily driven by the accelerated rate of decline in episodic and working memory and not by perceptual speed, visuospatial ability, and semantic memory (***Supplementary Table 14***). Conducting a sensitivity analysis with an alternative assessment of asymptomatic vs. symptomatic AD using the MMSE value^33^ indicated that the fast cluster exhibited odds of 2.8 times higher for symptomatic presentation than the slow group (***Supplementary Fig 9***). Furthermore, modelling using a random forest model trained on AD bulk modules to predict pseudotime in the primary cohort predicted higher pseudotime values in the fast group compared to the slow group (***Fig 5 c***). Feature importance analysis revealed that this prediction was predominantly driven by the neuronal injury response (F11_DOWN_ME2) and reactive astrocyte modules (F14_UP_ME2) (***Fig 5 d, Supplementary Fig 10***). We found statistically significant differences in module eigenvalues between the two groups, with the fast group exhibiting higher glial activity (***Fig 5 e, two upper panels***) and more pronounced neuronal injury response (***Fig 5 e, lower panel***). Projecting the genes and proteins up/down-regulated in the fast versus the slow group (***Supplementary Table 15***) to the discovery cohort snRNA dataset highlighted a high expression in Micro2, Astro0, and RORB^+^ excitatory neuron subtypes (i.e. Exc L3 RORB-CARTPT, Exc L4-6 RORB-LCN15) (***Fig 5 f***).

**Figure 5.**
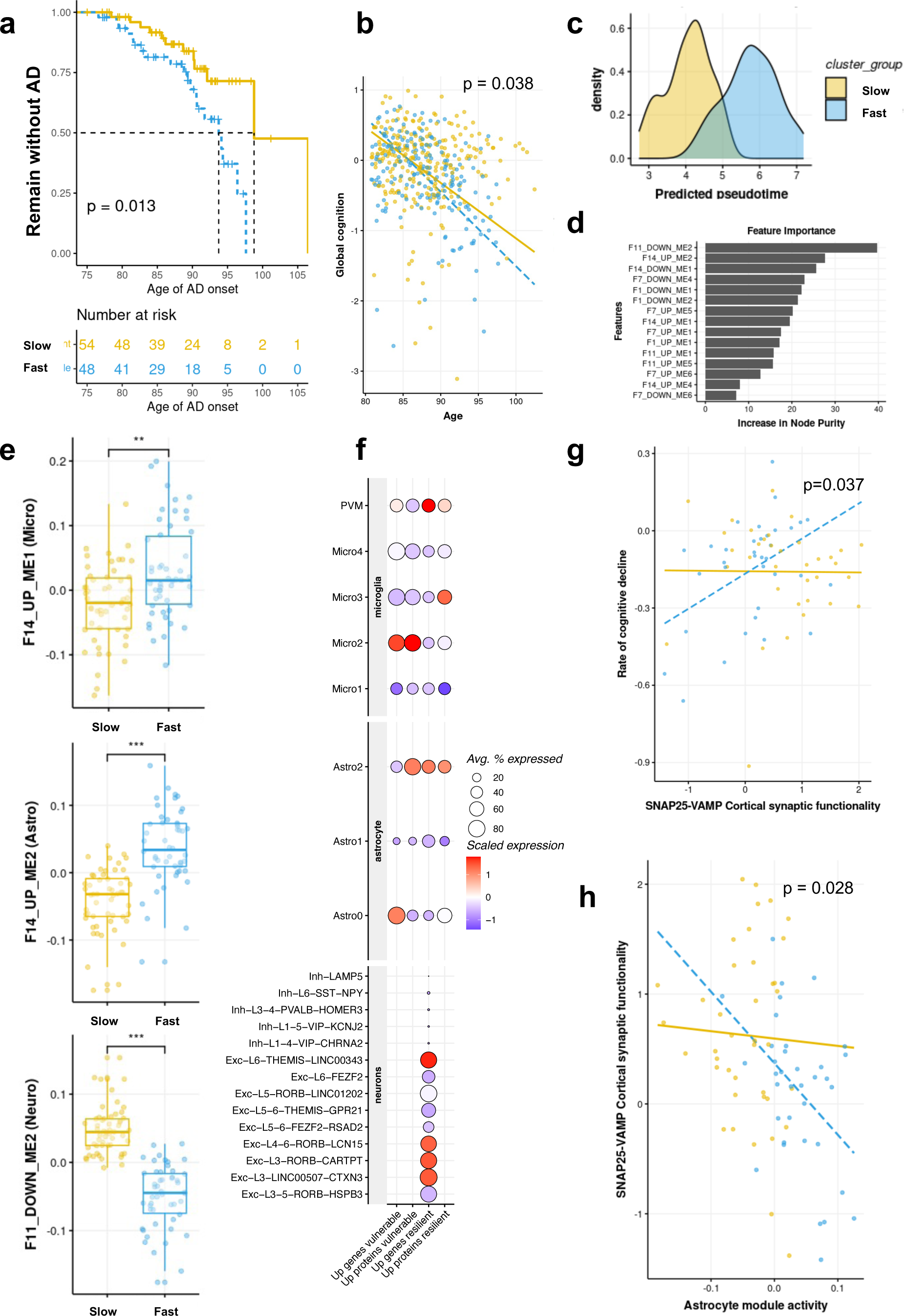
Identified glial activation mechanisms are related to the trajectory of cognitive decline and synapse dysfunction. **a)** Kaplan-Meier curves represent a delay in AD onset. Survival is quantified using longitudinal cognitive assessments, in which non-cognitive and mild-cognitive individuals without an AD diagnosis are considered. The analysis is stratified by subgroups identified in multi-omics clustering analysis and identifies fast (blue) and slow (yellow) groups based on their differential cognitive loss profiles. **b)** Linear mixed model assessing loss of global cognition as a function of age, highlighting a significant differential decline over time. **c)** Density plot depicting the predicted pseudotime in early AD, comparing slow and fast groups. The random forest model was trained on the discovery cohort AD-correlated bulk modules, and the trained model was applied to the validation cohort to predict pseudotime. **d)** A bar chart showing the importance of bulk modules, ranked by increasing node purity, in predicting pseudotime. **e)** Box plots comparing module scores between slow and fast patients. (Wilcoxon rank-sum test, adjusted p, *p≤0.05, **p≤0.01, ***p≤0.001). **f)** Dot plots showing the expression of up and down-regulated genes and proteins between fast versus slow groups (**Supplementary Table 15**) across microglial subtypes, astrocyte subtypes, and neuronal subtypes, with size representing the average percentage expressed and colour intensity denoting the difference in module score between Fast versus Slow groups**. g)** Significant interaction between SNAP-VAMP25 synaptic functionality and the rate of cognitive decline **h)** Synaptic functionality as a function of neurotoxic astrocyte module activity suggests a negative relationship only in the fast group.

To test for synaptic differences between the groups, we leveraged a SNARE proteomics dataset acquired on the same ROSMAP participants, quantifying synaptic protein-protein interaction of presynaptic components involved in the SNARE complex (known to mediate exocytosis) via heterologous capture ELISA^34^. We found that the level of SNAP25-VAMP interaction was diminished in the faster-progressing group (p<0.05). This reduction was not paralleled in other synaptic components, highlighting a specific dysfunction in SNAP25-VAMP interaction in the fast group (***Supplementary Table 16***). A linear mixed-effects model demonstrated an interaction between SNAP25-VAMP functionality and the slow/fast group classification, indicating that the fast group exhibits a more pronounced decline associated with SNAP25-VAMP dysfunction compared to the slow-progressing group (***Fig 5 g, Supplementary Table 19***). We found that the expression of the neuronal injury response module, F11_DOWN_ME2, was strongly associated with SNAP25-VAMP interactions, highlighting that the module captures variation related to synaptic dysfunction in AD (p=0.004, ***Supplementary Table 17***). SNAP25-VAMP interaction was also associated with the neurotoxic astrocyte module in the fast group but not with the microglia module (***Fig 5 h, Supplementary tables 19-20***). These findings highlight an association between increased astrocyte activation, synaptic dysfunction, and the rate of cognitive decline.

### Biomarkers in plasma and CSF: identifying groups at higher risk for more rapid cognitive decline

We investigated module proteins in CSF and plasma to find biomarkers and identify individuals at high risk for rapid cognitive decline in AD. We used plasma and CSF proteomics modules identified in a previous study from Dammer et al.^35^ and tested overlap with our candidate modules using Fisher’s exact tests. Module nomenclature and annotations are taken from the original study^35^ and details can be found in the methods section. In CSF, biomarkers were identified for microglial activation and neuronal injury response modules but not for the reactive astrocyte module (***Fig 6***). Notably, the overlap between Dammer’s CSF microglial modules M7 and F14_UP_ME1 includes proteins such as C3, CLEC7A, CD163, VPS26A, and VSIG4. For synaptic dysfunction, there was a significant enrichment of F11_DOWN_ME1 in both M6 and M8, with GAP43, STMN2, and NEFL being the overlapping proteins. These synaptic biomarkers correlated positively with pTau and showed up-regulation in AD CSF versus control. In plasma, biomarkers for microglial activation predominantly involve the complement system, with modules M28 and F14_UP_ME1 showing significant enrichment for the complement proteins C3, C1QA, C1QB, and C1QC. SNAP25 was detected in plasma as part of the M2 module, although the enrichment test with the neuronal injury module did not reach significance.

**Figure 6.**
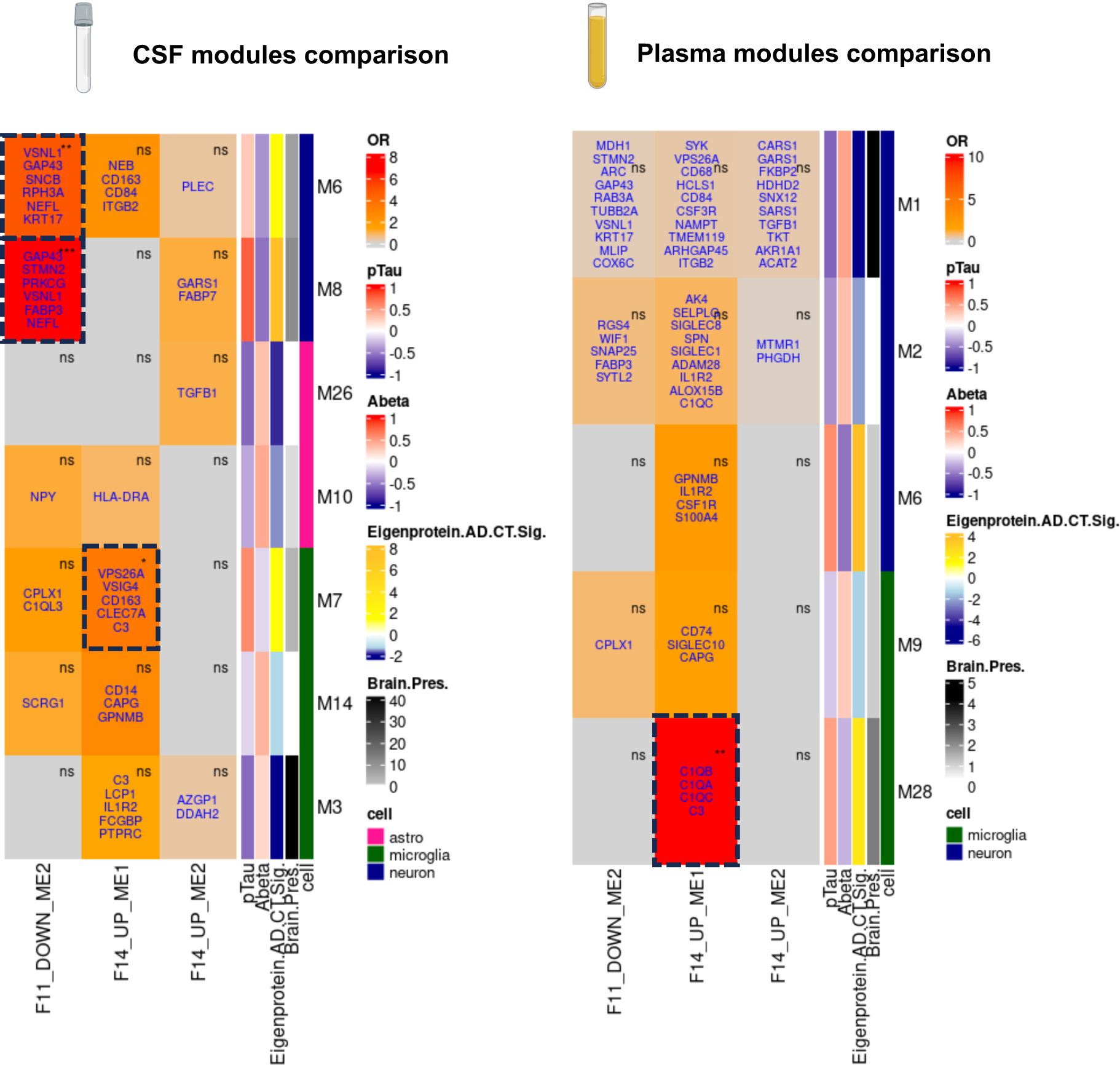
Biomarkers of glial activation and synapse dysfunction in CSF and plasma as potential biomarkers for Alzheimer’s accelerated progression. Reference CSF and plasma proteomics co-expression modules were taken from Dammer et al.^35^. Modules that were significantly differentially expressed in AD versus control and enriched for astrocytes or microglia and defined as being preserved in the brain were kept for analysis. OR: Fisher’s Exact test Odds Ratio. (Fisher’s exact test, *p≤0.05, **p≤0.01, ***p≤0.001). Gene names in blue overlap with the Dammer et al. modules compared to this study. All the row annotations were taken from Dammer et al. ^35^. pTau and Abeta show Pearson’s correlation values with the module eigenvalue. Brain Pres show brain preservation -log_10_(p-value). Eigenprotein AD.CT.sig shows - log_10_(p-value) of module eigenvalue in AD vs. Controls. Cell colour coding shows significant module enrichment for astrocytes, microglia, or neurons (p-value<0.05).

## Discussion

Whilst synaptic loss is a known correlate of cognitive decline in AD, the timing and role of glial activation in cognitive function remain unclear. This study used a multi-omics approach to investigate the molecular basis of AD, focusing on the dynamics of glial activation and its relationship with synaptic dysfunction and cognitive decline. We identified transcriptomic-proteomic modules linked to activated microglia and reactive astrocytes enriched in AD risk genes. By mapping these findings to single-nuclei data, we uncovered cell state-specific mechanisms involving microglial DAM/HLA profiles and neurotoxic astrocyte states. Early microglial activation was found to precede neuropathology, while later astrocytic responses were linked to cognitive decline and increased neuropathological burden. Our analysis shows individuals with rapid cognitive decline exhibit higher neurotoxic astrocyte reactivity, reduced SNAP25-VAMP interactions, and neuronal injury. These findings connect glial activation to synaptic dysfunction and cognitive decline in AD, highlighting roles for microglial activation in the genesis of AD and later astrocyte activation as a potential determinant of clinical symptom progression.

Prior large-scale multi-omics analyses have identified complex processes involved in AD pathogenesis^6,36–41^ and linked them to clinical and neuropathological traits, including changes during the asymptomatic to symptomatic phase^33^. Our unique contribution lies in identifying specific transcriptomics-proteomics modules (F14_UP_ME1, F14_UP_ME2, F11_DOWN_ME2) representing microglia, astrocytes, and neuronal injury mechanisms. We leveraged a novel approach to study cross-sectional *post mortem* data through pseudotime analysis, which leverages unsupervised integration of different brain regions (MTG, SOM) affected at various disease stages^42^. We build on previous single-cell studies identifying glial subpopulations involved in AD^11,43^, validating the cell-state specificity of AD-correlated glial activation and clarifying which subpopulations may mediate AD genetic risk. Subpopulations such as Micro2, enriched in HLA/DAM genes, and Astro0, enriched in neurotoxic astrocyte genes, were characterised. Contrary to findings suggesting a protective role for the microglia HLA state^9^, our data indicate otherwise, possibly due to the mixed DAM/HLA composition in our Micro2 subpopulation. We emphasised the distinct contributions of transcriptomics and proteomics in mediating AD genetic risk in microglia and astrocytes, with transcriptomics revealing numerous transcription factors regulating the microglial network (CEBPB, STAT1), while the astrocyte network may predominantly be regulated post-transcriptionally.

Previous research shows AD PRS is linked to cognitive decline rates, mainly through Aβ pathology^44^. Our findings align with this, demonstrating direct and indirect effects of pathway-specific PRS on AD progression. Indeed, we provide evidence for genetic risk being a determinant of microglial activation in AD, corroborating previous research^45,46^. By contrast, we show that astrocytic activation responses with AD, which appear to have a lower genetic association, are related to local environmental signals. This aligns with iPSC findings showing *TREM2*^R47H^ promotes proinflammatory microglia^15^, whereas *APOE*-ε4 increases Aβ aggregates and tau hyperphosphorylation alongside *APOE*-ε4 astrocytes with impaired Aβ uptake and cholesterol accumulation^47^.

We focused our differential cognitive decline analysis on Braak stages III-IV, considered a suitable therapeutic window for disease-modifying treatments as this phase marks the onset of clinical symptoms ^48^. We identified two early AD subgroups: one with earlier onset, faster cognitive decline, and higher odds of cognitive deficits, and another with slower cognitive decline. This aligns with previous studies uncovering heterogeneous cognitive trajectories in MCI ^49^. Furthermore, we linked this accelerated decline to a specific neurotoxic astrocytic response and neuronal injury response predominantly found in RORB^+^ excitatory neurons, suggesting a particular susceptibility corroborating previous research ^27,50^.

We then sought to link this differential cognitive to potential synaptic differences. Synaptic changes and SNARE transcriptional failure are associated with cognitive deficits^51,52^ and the concept of “cognitive reserve”, where higher baseline synaptic health helps resist neuropathological insults^34^. We observed a steeper relationship between the rate of cognitive decline and SNAP25-VAMP synaptic interaction in the fast-progressing group, which suggests that synaptic dysfunction may play a critical role in the accelerated deterioration observed in this subgroup. The association of SNAP25-VAMP interaction with the neurotoxic astrocyte module, but not with the microglia module, further implicates astrocytic involvement in this process. Given astrocytes’ essential role in synaptic plasticity and maintenance, the neurotoxic astrocyte state may exacerbate SNAP25-VAMP synaptic dysfunction. Our study establishes the translational relevance of these findings by identifying overlapping molecular signatures in CSF and plasma to serve as biomarkers for identifying individuals at risk of faster cognitive decline. Many of these biomarkers corroborate previous research, including SNAP25^53^, neurofilament light chain^54^ (NFL), GAP43^55^, or complement proteins^56^.

A limitation of this study is the limited proteome depth (∼3,000 proteins) compared to transcriptomics (∼20,000 transcripts), which likely resulted in missing proteomic changes. Similarly, the removal of the least explanatory transcripts to achieve a balanced MOFA integration may have led to missing important transcripts. Altogether, the MOFA factors and derived AD-correlated modules may have overlooked critical genes/proteins and pathways involved in the disease process. Another limitation of this study is the analysis of cross-sectional post-mortem data via pseudotime, which represents snapshots rather than continuous progression. To mitigate this and ensure our observations reflect dynamic disease processes, we related AD-correlated modules to longitudinal cognitive data from an independent cohort in early AD. To address potential survivor bias, we restricted the subset to those with follow-ups of 12 years or less. Furthermore, differences in population representation (ROSMAP early AD cohort mean of 89 years old vs. US average 83.7 years old^57^) limit the generalisability of the findings.

Overall, our integrative multi-omics study connects glial activation to synaptic dysfunction and cognitive decline in AD. Future research should apply integrative omics approaches prospectively and longitudinally, utilising CSF and plasma biomarkers to confirm glial activation dynamics and their effects on cognitive decline. The identified CSF and plasma biomarkers hold significant potential for stratifying rapidly progressing cases and identifying disease trajectories, warranting further exploration. Synaptic multi-omics measurements will enable the investigation of the mechanisms involving neurotoxic astrocytes and their potential role in SNARE synaptic dysfunction, providing insights into how these processes might accelerate synapse loss. Taken together, these would allow for a more personalised approach in the management and treatment of AD.

## Methods

### Brain tissue

This study was carried out in accordance with the Regional Ethics Committee and Imperial College Use of Human Tissue guidelines. Cases were selected based first on neuropathological diagnosis (NDC or AD) from UK brain banks (London Neurodegeneration [King’s College London], Newcastle Brain Tissue Resource, Manchester Brain Bank, South West Dementia Brain Bank [Bristol], and Parkinson’s UK [Imperial College London] Brain Bank). MTG and SOM cortex from a final set of 56 donors, including 23 controls (Braak stage 0–II) without evidence of clinically significant brain disease (non-diseased controls, NDC) and 33 AD cases (Braak stage III-VI) were used (total of n=112 brain samples). Cortical samples from two regions were prepared from each brain to characterise pathology, transcript expression and protein abundance.

### Bulk transcriptomics

Samples were randomized for RNA extraction to reduce batch effects between conditions. Trizol was added to homogenise tissue, followed by Chloroform. Rigorous vortexing of the samples resulted in homogenisation. A Qiagen RNeasy micro kit (cat no 74004) was used for extraction and purification as per the manufacturer’s instructions. Briefly, samples in MinElute Spin Columns were treated sequentially with buffers RW1 and RPE. Samples underwent DNase I digestion, and the resulting RNA was eluted in RNase-free water. Samples were quantified using the nanodrop, and RIN values were obtained using an Agilent Bioanalyser (RNA nano chip and reagents cat no 5067-1512).

Samples were normalised for concentration and underwent ribodepletion using the New England Biolabs rRNA depletion kit (cat no E7400X). Genomic Libraries were constructed using the New England Biolabs Library construction kit (cat no E7760L), using the NEBNext Multiplex oligos for Illumina (cat no E6442S). Libraries were quantified using a Qubit fluorometer and pooled for sequencing using a HiSeq4000 at 50 million reads per sample. Samples were split across multiple lanes to avoid batch effects. The bulk RNA-seq data was pre-processed using the nextflow pipeline nf-core-rnaseq v3.10.1 using default parameters with alignment and quantification performed by the included STAR and RSEM tools. The *post mortem* tissue samples were rejected during quality control if less than 90% of reads could be aligned with a minimum requirement of 30 million aligned reads.

### Bulk proteomics and pre-processing

Tissue samples had been stored in different amounts of optimal-cutting-temperature (OCT) solution. To remove this and standardise tissue preparation, all samples were cryo-sectioned into two 80μm sections, and OCT was removed as previously described^58^. Briefly, 1mL of 70% (V/V) ice-cold ethanol was added to the sections with end-over-end rotation for 2 minutes. The ethanol was removed by centrifuging for 2 minutes at 10,000xg at 4°C, and the wash was repeated for a total of 5 washes. Washes were repeated as described, but with water (4 washes) and then 50mM ammonium bicarbonate (3 washes) before allowing the sections to air-dry for 5 minutes. Protein was extracted with 50μL 8M urea in 100mM ammonium bicarbonate with the addition of 0.1mM phenylmethylsulfonyl fluoride (PMSF) protease inhibitor. Lysis was ensured with a probe-sonicator with 10x1second pulses on 30% amplitude, with samples on ice throughout. Lysates were clarified through centrifuging at 10,000xg for 10 minutes, 4°C and the supernatants frozen at -80°C until all protein extraction was completed.

Following a protein assay (DC Protein Assay, BioRad), 20μg of protein was reduced, alkylated and digested using an in-solution digestion approach automated using an Andrews^+^ pipetting robot with a 96-PCR Plate Peltier+ (Waters). Briefly, at 4°C, 20μg of protein was combined with water and a pre-mixed digestion solution, resulting in 0.2μg trypsin (Sequencing Grade Modified Trypsin, Promega), 100mM ammonium bicarbonate, 40mM chloroacetamide and 10mM Tris(2-carboxyethyl)phosphene and then heated to 37°C for 16 hours before being chilled to 4°C and acidified to 0.5% (V/V) trifluoro-acetic acid (TFA). Peptides were desalted in a semi-automated manner using an Andrews+ pipetting robot in combination with a Vacuum+ (Waters) by solid-phase extraction on an Oasis HLB μElution Plate (Waters). Briefly, the plate was pre-equilibrated by washing three times with 100μL 0.1% (V/V) TFA. All samples were added to the plate and washed a further 3 times with 100μL 0.1% (V/V) TFA because being eluted with 50μL followed by 100μL 60% acetonitrile into 96-well QuanRecovery plates (Waters) and dried at 60°C using a Speedyvac (Eppendorf). All volumes were drawn through the plate by negative pressure. All samples were reconstituted into 200μL 0.1% formic acid (FA) by sonicating for 10 minutes in a water bath. A pooled sample was generated from 20μL of all samples and used as a study reference (SR) and for spectral-library generation.

For spectral library generation, 100μg of protein was fractionated into 11 using high-pH reversed phase fractionation kit (Pierce, ThermoFisher) and analysed by LC-MS/MS. Peptide separation was performed using a 0.3x150mm Kinetex XB-C18 column (Phenomonex) in-line with an M-Class HPLC (Waters) using top 60 data-dependent acquisition on a 7600 ZenoTOF (Sciex). Each fraction was injected twice with data acquired over a precursor mass range of 400 to 655mz or 650 to 905mz. Precusors were accumulated for 0.1s with MSMS accumulation set to 0.01s and data acquired across 140-1800mz. Sampled ions were excluded for 6 seconds after a single acquisition with priority given to ions between +2 and +4 charge with greater than 300 counts per second. The peptides were eluted from the LC by increasing the proportion of buffer B (Acetonitrile + 0.1% (V/V) FA) from 3% to 30% compared to buffer A (Water + 0.1% FA) over 44 minutes. B was increased to 80% over the following 2 minutes and held for 2 minutes before returning to 3% for 5 minutes. The flow rate was set to 5μL/minute throughout.

Individual brain samples were analysed by LC-MSMS on the same instrumentation using zenoSWATH data-independent acquisition (DIA) with 85 variable windows. MS was acquired between 400-1500*mz* with an accumulation time of 0.1s. Over 85 variable windows spanning 399.5 to 903.5*mz*, MSMS was acquired between 140-1800*mz* using dynamic collision energy ranging from 19 to 43V. Peptides were separated on a 0.3x150mm Kinetex XB-C18 column (Phenomonex) with solvent B increasing from 3 to 30% over 20 minutes before increasing to 80% over 1 minute for a two-minute wash. The column was re-equilibrated to 3% B for 5 minutes before triggering the next sample injection. The flow rate was 5μL/minute throughout and 3μL of sample was injected in partial-loop mode. The instrument was automatically calibrated every 6 injections, which constitutes 5 samples and 1 SR. All instrumentation was operated through OS version 3.1 (Sciex).

Data-dependent acquisition data was analysed in Fragpipe^59^ (version 19) by searching the human SwissProt canonical database downloaded on the 08/09/2022 using defaults. Carbamidomethylation of cysteine was specified as a fixed modification with variable oxidation on methionine. Mass tolerance was +/- 20ppm, and N-terminal methionine was removed. Precursors with a charge between 1 and 4 were allowed with a minimum length of 7 amino acids and a maximum of 2 missed cleavages. Data was exported as a spectral library with FDR specified < 0.01 containing 4636 protein groups and 51105 precursors.

DIA data was searched using DIA-NN^60^ (version 1.8.1) using the spectral library described above. Match-between runs was enabled, and DIANN was allowed to determine optimum values for scan width (6), MS (13.6ppm) and MSMS (14.2ppm) tolerance. Outputs were filtered to 0.01 FDR.

### Single nuclei transcriptomics

This data comes from the preprint from our group ^26^, whose samples partly overlapped with the bulk cohort (42 out of 112 samples). Single nucleus isolation was performed on 90 tissue blocks from the MTG and SOM regions (n = 26, NDC; n = 42, AD), following established protocols^61,62^. Approximately 7,000 isolated nuclei per sample were processed and sequenced using the 10x Genomics Chromium Single Cell 3’ platform. Pre-processing, quality control, integration, clustering, and cell type assignment of snRNA sequencing data were conducted as described in^63^. The present analysis focuses on astrocyte, microglia, and neuronal subsets. Methods for the sub-clustering of neuronal sub-populations are detailed in^27^. Sub-clustering of astrocytes and microglia followed the same steps and used a resolution of 0.1 for astrocytes and 0.2 for microglia.

### Immunohistochemistry (IHC)

IHC was performed on FFPE sections from the MTG and SOM of each brain studied and paired with material from the cryopreserved contralateral hemisphere used for nuclear preparations for snRNA sequencing. Standard immunostaining procedures, as recommended by the manufacturer, were followed using the ImmPRESS Polymer (Vector Laboratories) and Super Sensitive Polymer-HRP (Biogenex) kits. After dewaxing and rehydration of slides, endogenous peroxidase activity was blocked with 0.3% H_2_O_2_, followed by antigen retrieval (***Table 1***). For immunostaining using ImmPRESS kits, non-specific binding was blocked using 10% normal horse serum. Primary antibodies were incubated overnight at 4 °C. Species-specific ImmPRESS or Super Sensitive kits, and DAB were used for antibody visualisation. Tissue was counter-stained by incubation in Mayer’s haematoxylin (TCS Biosciences) for 2 min, followed by dehydration, clearing and mounting.

**Table 1:** Antibodies used for Immunohistochemistry. CB=citrate buffer. EDTA= ethylenediaminetetraacetic acid.

| Antibody | Supplier | Dilution | Antigen retrieval | Staining kit |
| --- | --- | --- | --- | --- |
| **β-Amyloid (4G8)** | Biologend, 800711 | 1:15,000 | Steamer, CB pH6 | Super Sensitive Polymer-HRP |
| **pTau (PHF-1)** | The Feinstein Institutes for Medical Research | 1:5000 | Steamer, CB pH6 | Super Sensitive Polymer-HRP |

### Processing of bulk transcriptomics-proteomics data

Bulk transcriptomics and proteomics data were processed using the Omix R package V1.0^64^ to generate a comprehensive multi-assay dataset, with parameters specified in supplementary table 2. After filtering, 20231 genes and 2945 proteins were kept for analysis in the transcriptomics and proteomics data, respectively. Variations related to sex, *post mortem* delay, and batch effects were regressed out to minimise confounding factors. A quality control (QC) report, including all processing parameters and steps, is provided in ***Supplementary File 1***.

External validation of disease modules was sought in two independent large-scale AD cohorts with publicly available bulk transcriptomics and proteomics data (ROSMAP and MSBB). The same parameters described above were used to process these datasets.

### Bulk multi-omics integration

Integration of the bulk multi-omics data was performed using the Multi-Omics Factor Analysis (MOFA) model^23^. Prior to integration, scaling of the different modalities was carried out to ensure comparability. For the transcriptomics data, the 2,945 most variable features were selected to achieve balanced views between the transcriptomics and proteomics modalities. This subsetting was crucial to harmonise the data and enhance the robustness of the integration process. Other parameters were kept to default values.

### Cross-sectional pseudotime trajectory

To compensate for the lack of longitudinal tissue data in *post mortem* samples, we employed a pseudotime trajectory analysis to model transcriptomic-proteomic dynamics across a disease continuum. It uses the same concept as in pseudotime inference with scRNA-seq^65^, except here, each data point is a bulk sample derived from a patient, and pseudotime represents an inferred trajectory along which patient samples are ordered, modelling disease progression from early to advanced stages. The disease trajectory was derived using the slingshot algorithm^66^ on a two-dimensional Uniform Manifold Approximation and Projection (UMAP) representation of AD-relevant and brain region-specific MOFA factors. Samples were ranked according to their positions within the inferred trajectory and respective Braak stages. Samples were ranked within tiers of high variability to simulate Braak stages. This step involved categorising samples into early (Braak 0-II), mid (Braak III-IV), and late (Braak V-VI) stages based on pathological markers and clinical observations. The final pseudotime values were determined by fitting a linear model between the derived disease trajectory and the sample ranks within each Braak tier. This provided a continuous measure of disease progression (***Fig 1b-c***). This method allows us to model AD progression as a continuous process, positioning each patient along an inferred gradient of disease severity.

### Transcriptomics-proteomics co-expression modules

In MOFA, feature (gene) weights from factors 1, 7, 11, and 14 strongly correlated with AD pathological variables. Features with weights exceeding ±1.5 SD from the mean were used to define the signature. Pairwise correlations between features in the multi-omics signature were assessed, and a multi-omics network was constructed where feature pairs showed a correlation exceeding a user-defined threshold (default=0.3). This network was then dissected into functionally relevant modules using community detection algorithms like Louvain^67^, resulting in “disease-relevant modules” that aid in understanding disease mechanisms. Module eigenvalues were calculated via Principal Component Analysis on scaled transcriptomics and proteomics expression data, and the first principal component was assigned as the module eigenvalue. Modules identified in the primary cohort were projected onto independent datasets using the same features and eigenvalue computation method.

### Differential module analysis

Differential module analysis identified genes with significant expression differences between case and control groups. Linear models using limma (v.3.50.3)^68^ were fit for each module to assess differential module changes against (1) pseudotime, (2) AD vs. controls, and adjusting for age, sex, PMI, brain region, *TREM2 and APOE* genotypes, and noise factors (factors not related to AD variation). P-values from the moderated t-tests were adjusted for multiple testing using the Benjamini-Hochberg procedure to control the false discovery rate^69^. Modules were prioritised based on their significance levels, selecting those with the most significant differential expression for further analysis.

### Module cell type enrichment

Cell-type enrichment analyses were conducted using Expression-Weighted Cell Type Enrichment (EWCE) (v1.2)^24^. This analysis utilised a cell-type data reference from the Allen Human Brain Atlas^25^ to determine the enrichment of specific cell types within the identified modules (50277 genes across 9 cell types).

### Module functional enrichment

Functional enrichment analysis was performed using Enrichr^70^. This streamlined pathway analysis allowed for the generation of pathway reports utilising common pathway databases such as KEGG, Reactome, BioCarta, MSigDB, and Gene Ontology. The results were presented using our internally developed, intuitive, and interactive network visualisations^64^ (***Supplementary Files 2-9***).

### Projecting bulk modules and gene sets in single nuclei data

To project gene modules identified from bulk RNA sequencing data onto single-cell RNA-seq data, we utilised the hdWGCNA package^71^. First, we generated single-cell gene expression data in a Seurat object containing normalised and scaled expression matrices. We then identified the single-cell equivalents of the bulk RNA-seq gene modules using the *project_bulk_modules* function from hdWGCNA. This function uses a signed network approach to calculate module eigengenes from the bulk RNA-seq data and projects them into the single-cell RNA-seq space by computing correlations with single-cell expression profiles. The projected module scores provide a measure of module activity in individual cells. We subsequently visualised module scores across different cell types or conditions using UMAP, identifying cell types with significant module enrichment. This analysis enables the identification of cellular contexts driving module activity and provides insights into the cellular heterogeneity of gene module regulation.

We used the same approach to project the neuroprotective and neurotoxic gene set on the astrocyte subpopulations. Gene sets were obtained from *Cameron et al.*^13^, mapped to human genes and filtered for unique genes to avoid overlapping genes between neurotoxic and neuroprotective genes (***Supplementary Fig 6***a-b***, Supplementary Table 8)***. We also projected the microglia states defined in *Mancuso et al.*^9^ following the same approach (***Supplementary Fig 6***c).

### AD GWAS enrichment analysis

To perform genetic enrichment analysis of modules, we utilised MAGMA software (v1.10)^28^. First, we extracted SNP locations and p-values from harmonised summary statistics of the different GWAS studies ^29–31^. Using the SNP location data and an NCBI gene location file (NCBI38.gene.loc), we annotated SNPs to genes with *MAGMA’s -- annotate* command. We then used the annotated gene information to conduct a gene-based association analysis using reference genotypes from the 1000 Genomes Project (European population). The gene-based p-values were computed using the summary statistics file and the previously generated gene annotation file. To identify significant genetic associations across multiple studies, we performed a meta-analysis using gene-based association results from the three GWAS studies (Bellenguez_magma.genes.out, Janssen_magma.genes.out, and Wightman_magma.genes.out) and combined results using the *--meta* command to obtain meta-analysed p-values (meta_analysis.genes.out). For the genetic enrichment analysis of modules, we used the individual and combined gene-level statistics with *MAGMA’s --set-annot* command on a text file containing module entrez-id features.

### Transcription factor target gene enrichment analysis

Transcription factor enrichment analysis serves as a powerful tool to uncover the regulatory layers of the network. It focuses on identifying transcription factor (TF) proteins whose target RNAs are notably present within specific disease modules. This method leverages an external dataset of TF and their targets, generated from previous ChIP-seq experiment data^72^, and assesses whether the overlap between TF target genes and each disease module is statistically significant through Fisher’s exact test.

### Polygenic Risk Score

To compute polygenic risk scores (PRS), we used the PLINK software (v1.90)^73^ based on whole genome sequencing data from the ROSMAP cohort and GWAS summary statistics. The SNPs included in the scores were selected from genes identified as significant (p<0.05) in driving genetic enrichment within microglia and astrocyte functional modules, as determined in the AD GWAS enrichment step. To calculate the PRS, we applied PLINK’s scoring function to derive separate scores for astrocytes (calculated using effect sizes from the Wightman et al. GWAS) and microglia (calculated using effect sizes derived from our meta-analysis). Scores were computed across a range of predefined p-value thresholds from 10^-7^ to 0.5, after which both the astrocyte and microglia PRS were scaled to ensure comparability. The scaling process involved normalising the scores to a standard mean and variance across the cohort. To determine the optimal PRS for predicting phenotypic outcomes, we conducted regression analyses across various p-value thresholds (from 10^-7^ to 0.5) against two phenotypic variables: global pathology and activity of the microglia/astrocyte modules. The best-fit PRS was selected based on the model that explained the highest proportion of phenotypic variance. The optimal model fit for the Microglia PRS was achieved at a p-value threshold of 0.001, which included 68 SNPs. Of these, 61 SNPs were present in the ROSMAP genomic data. This model maximised the R² value when modelled against microglia module activity, indicating a strong predictive capability. The best fit for the Astrocyte PRS was found at a p-value threshold of 0.005, encompassing 71 SNPs, with 21 detectable within the ROSMAP genomic data. This model demonstrated the highest R² when correlated with global pathology measures. These findings are detailed further in **Supplementary** Figure 8 and ***Supplementary Tables 10and 11***, which include comprehensive lists of the SNPs used in each model and their respective contributions to the PRS calculations.

### Rate of cognitive decline

To assess the rate of cognitive decline, we computed participant-specific rates over a 3-year interval. The data were grouped by participants, and the rate of cognitive decline was measured by calculating the difference in cognitive scores between the most recent time point (i.e., the time of death) and the closest available follow-up year to 3 years before death. This individual rate measures the speed of cognitive decline over time, where a negative rate indicates a decline in cognition, while a positive rate indicates an improvement. We restricted the time window to be close to the time of death to homogenise the varying follow-up timing in the ROSMAP cohort and to ensure that these rates of cognitive decline accurately reflect brain changes quantified *post mortem*.

Of the 132 participants, 126 had available longitudinal measurements, with at least 3 measurements per participant. For 90.5% (114/126) of the participants, these measurements spanned a true 3-year interval. In a few cases, measurements were taken at intervals closest to the 3-year span, specifically 2-year (6.35%, 8/126), 4-year (2.38%, 3/126), and 5-year (0.794%, 1/126) intervals. To account for these varying time intervals, we standardised the rate of cognitive decline by dividing the score difference by the corresponding time interval. This standardisation ensured that the computed rate accurately reflected the speed of cognitive change, regardless of the exact measurement interval.

### Multi-omics subtyping of early AD

We employed iClusterBayes from the iClusterPlus R package (v1.22.0)^32^ to cluster AD participants. First, we optimised the number of clusters within the early AD ROSMAP subset (n=132) using the Cluster Prediction Index (CPI) from the IntNMF package (v1.2)^74^ and the Gap-statistics from the MOGSA package (v1.28)^75^. Data pre-processing included shifting the values of each dataset to be non-negative and rescaling them to the range [0, 1] to meet the non-negativity constraint of IntNMF. Using the *nmf.opt.k* function from the IntNMF package, we calculated the CPI by performing non-negative matrix factorisation (NMF) across a range of cluster numbers, providing a score for each number of clusters tested. Concurrently, the Gap-statistics method, implemented via the *moGap* function from the MoCluster package, compared the within-cluster dispersion against a null reference distribution, producing a Gap statistic for each number of clusters. The optimal number of clusters of two was determined by combining the CPI and Gap-statistics results, selecting the number that maximised the sum of the CPI score and Gap statistic. Next, we ran a Bayesian integrative clustering analysis using the *iClusterBayes(type=c(“gaussian”, “gaussian”), K=1*) function on transcriptomics and proteomics to identify common underlying structures. The analysis involved 18,000 burn-in iterations and 12,000 drawing iterations, with a thinning parameter of three to reduce autocorrelation. A prior probability parameter of 0.5, a standard deviation of 0.5, and a beta variance scaling factor of one were used. The posterior probability cutoff for cluster membership assignment was set at 0.5.

### Differential cluster analysis and pathways enrichment

Differential cluster analysis was performed using the *clustering_DE_analysis* function from the Omix package. Covariates included PMI, age, and sex. A log2 fold change threshold of 0.1 was applied for proteomics and 0.5 for transcriptomics. Comparisons were made between cluster sets CS2 and CS1, representing fast versus slow groups, respectively. Multiple testing was adjusted using the Benjamini-Hochberg procedure. For pathway enrichment analysis, Enrichr was used to generate pathway reports. The analysis included several pathway databases: GO Molecular Function 2021, GO Cellular Component 2021, GO Biological Process 2021, Reactome 2016, KEGG 2021 Human, MSigDB Hallmark 2020, and BioCarta 2016. These reports provided insights into the functional implications of the differential expression findings.

### Kaplan-Meier analysis

We used the Kaplan-Meier method to analyse the delay in clinical AD onset among participants classified into two cluster groups. The analysis was conducted using the *survfit* function from the survival package (v3.2-3) in R. Participants were considered event-free until the clinical onset of AD (Clinical cognitive diagnosis summary, dcfdx ≥ 4 [*Clinical cognitive diagnosis summary]*). The follow-up period was capped at 12 years, based on our findings that longer follow-up times skewed survival estimates and thereby caused survivor bias (n= 102/132, ***Supplementary Fig 9***). Individuals who never experienced AD onset were censored at the time of their last follow-up (death). We compared the survival curves of the two cluster groups, focusing on the time to AD onset, with curves visualised using the *ggsurvplot* function, incorporating confidence intervals and risk tables for detailed comparison.

### Random forest model to predict pseudotime

The internal dataset was partitioned into training (80%) and testing (20%) using the *CreateDataPartition* (caret package in R [v.6.0.94]) to ensure that the distribution of the outcome (pseudotime) is preserved in both the training and testing sets. We then trained a regression-based random forest model with the 15 significant transcriptomics-proteomics modules as predictors and pseudotime as the target variable, utilising feature importance computation to identify key predictors. Hyperparameter tuning was performed using a grid search over mtry (1-15) and ntree (1000-2500) values within a 10-fold cross-validation, with Root Mean Squared Error (RMSE) as the performance metric. The optimal hyperparameters (mtry = 3, ntree = 1500) were identified and used to train the final model on the entire training dataset (***Supplementary Fig 10)***. The final model performance was evaluated on the test set, yielding an RMSE of 1.105, R^2^ of 0.655, and MAE of 0.931. Variable importance was assessed using two metrics: the percent increase in mean squared error (%IncMSE) and the increase in node purity (IncNodePurity), providing insight into the key biological processes underlying pseudotime variation. The trained random forest model was applied to predict pseudotime on the external early AD ROSMAP dataset using the same projected modules initially identified in the internal dataset.

### SNARE proteomics

Quantitative immunoassay studies are all performed in the Honer Lab at the University of British Columbia (Vancouver BC, Canada). Detailed ELISA methodologies were published earlier^34^. 77 out of the 132 early ROSMAP participants sub-cohort have synaptic measurements for SNAP24-VAMP functionality in the combined cortical regions (*synap_3cort_capture4 variable*, n=40/70 slow and n=36/62 fast)

### Linear mixed-effects model with interaction terms

**Interaction Models for Cognitive Trajectories (*Supplementary Table 14*):**

We employed a linear mixed-effects model using the *lmer* function from the lme4 package (v.1.1)^76^ to evaluate the interaction between age and clusters on cognitive trajectories. The dependent variables were global cognition scores (cogn_global, cogn_ep, cogn_ps, cogn_po, cogn_se, cogn_wo). Fixed effects in the model included interactions between age (year) and clusters (cluster_group), with covariates such as sex (msex), baseline age (age_bl), baseline cognition (cog_bl), and years of education (educ). A random intercept for each participant (projid) accounted for intra-individual variability. The age variable was derived by summing baseline age and follow-up year. An age threshold of 80.74 was identified at the cognitive level intersection between the two groups. Interaction effects were visualised using the *interact_plot* function from the interactions package.

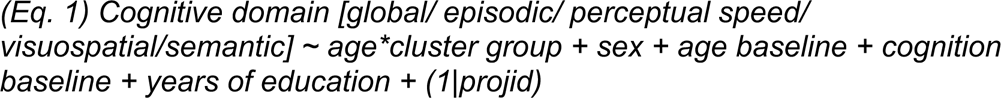

**Interaction Models for Synaptic Functionality (*Supplementary Tables 18-20*):**

To study the relationship between synaptic functionality and various factors, we used interaction models focusing on the SNAP25-VAMP synaptic complex (synap_3cort_capture4 variable). We analysed the following models:

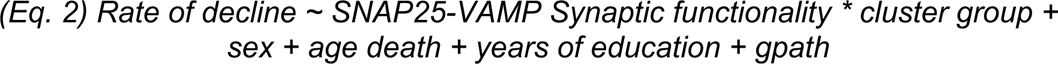

This model examines how the rate of cognitive decline preceding death relates to SNAP25-VAMP synaptic functionality and its interaction with cluster group, controlling for sex, age at death, years of education, and global pathology (gpath).

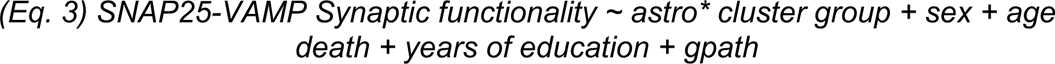

This model looks at the relationship between SNAP25-VAMP synaptic functionality and the astrocyte F14_UP_ME2 module, with an interaction term for cluster group, adjusting for the same covariates.

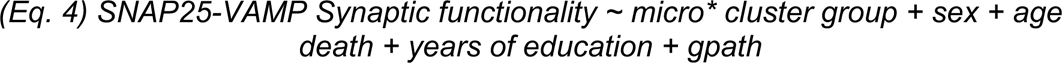

This model investigates the relationship between SNAP25-VAMP synaptic functionality and the microglia F14_UP_ME1 module, including an interaction term for cluster group and controlling for the same covariates.

### CSF and plasma proteomics modules overlap

CSF and plasma proteomics modules and relevant annotations were taken from a previous analysis by Dammer et al.^35^. We used Dammer’s modules, which showed significant enrichment of neuronal, astrocyte, or microglia cell types and were significantly differentially expressed in AD versus controls. The overlap between our modules and CSF and plasma proteomics modules was tested using Fisher’s exact test via the Gene Overlap package (v4.4). This test calculates a p-value to evaluate whether the observed overlap is likely if the gene sets are independent. It also provides an odds ratio, a measure of the association strength between the gene sets. An odds ratio of 1 or less indicates no association. We considered all expressed genes as the background set, following default settings.

## Data availability

### Internal cohort

#### UK DRI Multi-‘omics Atlas Project

This study’s metadata and patient IDs can be found in the following Synapse repository: syn61606248. The BrainBankNetworkID in the sample sheet must be cross-referenced with the omics datasets to ensure the inclusion of the same donors.

- Proteomics: syn53424292
- Transcriptomics: syn52943644
- Histopathology: syn52658344
- Single nuclei transcriptomics: syn53165790
  ○ Seurat objects of filtered microglia, astrocyte, and neuronal populations are available upon request.

### External cohorts

*Selected participants’ IDs and related derived features (modules, cluster groups, PRS scores, etc.) used in the current study can be found in the study Synapse repository syn61606248*

#### ROSMAP

- Transcriptomics and TMT proteomics data: Publicly available at Synapse under IDs syn4164376 and syn21971722, respectively.
- Whole genome sequencing: syn11707419
- SNARE Synaptic Proteomics: Data can be requested from [RADC Data Requests](https://www.radc.rush.edu/requests/data.htm).

#### MSBB

- Transcriptomics and TMT proteomics data: Publicly available at Synapse under IDs syn16796121 and syn24995077, respectively.

#### GWAS Studies

- MAGMA: cncr.nl/research/magma
- 1000 Genomes reference data: Summary statistics are available at cncr.nl/research/summary_statistics

#### CSF/Plasma Module Memberships from Dammer et al

- Supplementary information is available at https://doi.org/10.1186/s13195-022-01113-5

## Code Availability

All codes concerning the transcriptomics-proteomics integration and related downstream analyses is available as an R package that can be downloaded from the following GitHub repository: https://github.com/eleonore-schneeg/Omix. All codes for other downstream analyses are available upon direct contact with the lead author. The final random forest model to predict pseudotime is available from the following Synapse repository: syn61606248.

## Abbreviations

**A**

- AD: Alzheimer’s Disease
- Aβ: Amyloid Beta

**C**

- CDR: Clinical Dementia Rating
- CRM: Cytokine and Chemokine Response
- CSF: Cerebrospinal Fluid

**D**

- DAM: Disease Activated Microglia

**E**

- EWCE: Expression-Weighted Cell Type Enrichment

**G**

- GWAS: Genome-Wide Association Studies

**H**

- HLA: Antigen Response

**I**

- IRM: Interferon Response

**M**

- MCI: Mild Cognitive Impairment
- MMSE: Mini-Mental State Examination
- MOFA: Multi-Omics Factor Analysis
- MTG: Middle Temporal Gyrus

**N**

- NFT: Neurofibrillary Tangles

**P**

- PMI: Post-Mortem Interval
- PRS: Polygenic Risk Score

**S**

- scRNA: Single-cell RNA Sequencing
- SnRNA: Single-nucleus RNA Sequencing
- SNARE: Soluble N-ethylmaleimide-sensitive factor Attachment protein REceptor-mediated exocytosis
- SOM: Somatosensory Cortex

**T**

- TF: Transcription Factor
- TREM2: Triggering Receptor Expressed on Myeloid Cells 2

## Supporting information

Reports

## Data Availability

Internal cohort
UK DRI Multi-'omics Atlas Project
This study's metadata and patient IDs can be found in the following Synapse repository: syn61606248. The BrainBankNetworkID in the sample sheet must be cross-referenced with the omics datasets to ensure the inclusion of the same donors.
Proteomics: syn53424292
Transcriptomics: syn52943644
Histopathology: syn52658344
Single nuclei transcriptomics: syn53165790
Seurat objects of filtered microglia, astrocyte, and neuronal populations are available upon request.
External cohorts
Selected participants' IDs and related derived features (modules, cluster groups, PRS scores, etc.) used in the current study can be found in the study Synapse repository syn61606248
ROSMAP
Transcriptomics and TMT proteomics data: Publicly available at Synapse under IDs syn4164376 and syn21971722, respectively.
Whole genome sequencing: syn11707419
SNARE Synaptic Proteomics: Data can be requested from RADC Data Requests.
MSBB
Transcriptomics and TMT proteomics data: Publicly available at Synapse under IDs syn16796121 and syn24995077, respectively.

https://www.synapse.org/Synapse:syn61606248

https://www.synapse.org/Synapse:syn52658322

## Acknowledgements

We thank the donors and their families for the use of human brain tissue in this study and the UK brain bank staff for making it available. Tissue samples were provided by the London Neurodegenerative Diseases Brain Bank at King’s College London. The brain bank receives funding from the UK Medical Research Council and as part of the Brains for Dementia Research programme, jointly funded by Alzheimer’s Research UK and the Alzheimer’s Society. Tissue for this study was provided by the Newcastle Brain Tissue Resource which is founded in party by a grant from the UK Medical Research Council (G0400074), by NIHR Newcastle Biomedical Research Centre and Unit awarded to the Newcastle upon Tyne NHS Foundation Trust and Newcastle University, and as part of the Brains for Dementia Research Programme jointly funded by Alzheimer’s Research UK and Alzheimer’s Society. Tissue samples were supplied by The Manchester Brain Bank, which is part of the Brains for Dementia Research programme, jointly funded by Alzheimer’s Research UK and Alzheimer’s Society. We acknowledge the Oxford Brain Bank, supported by the Medical Research Council (MRC), the NIHR Oxford Biomedical Research Centre and the Brains for Dementia Research programme, jointly funded by Alzheimer’s Research UK and Alzheimer’s Society. Tissue samples and associated clinical and neuropathological data were supplied by Parkinson’s UK Brain Bank at Imperial, funded by Parkinson’s UK, a charity registered in England and Wales (258197) and in Scotland (SC037554). We are grateful to Diana P. Benitez for her support in the human tissue management.

Infrastructure, notably the LMS/NIHR Imperial Biomedical Research Centre Flow Cytometry Facility and the Imperial BRC Genomics Facility, was supported by the National Institute for Health Research (NIHR) Biomedical Research Centre (BRC). ST was supported by an “Early Postdoc Mobility” scholarship (P2GEP3_191446) from the Swiss National Science Foundation and a “Clinical Medicine Plus” scholarship from the Prof Dr Max Cloëtta Foundation (Zurich, Switzerland). JH acknowledges support from the Dolby Foundation. PMM acknowledges generous personal support from the Edmond J Safra Foundation and Lily Safra and an NIHR Senior Investigator Award. JSJ was supported by funding from Alzheimer’s Society (grant number 599 (AS-DRL-22-008)).This work was supported by the UK Dementia Research Institute, which receives funding from UK DRI Ltd., funded by the UK Medical Research Council, Alzheimer’s Society, and Alzheimer’s Research UK. The study is from the UK Dementia Institute Multi-omics Atlas Project for Alzheimer’s Disease (MAP-AD; map-ad.org). This study was partially supported by an investigator-initiated grant from Biogen IDEC to PMM and JSJ

## Author Contributions

E.S. performed data processing, bioinformatics, statistical analyses and interpretation.

E.S. also prepared the figures, drafted the manuscript, and was responsible for all subsequent revisions and corrections.

N.F Provided guidance on data analysis and interpretation

M.T Performed bulk transcriptomics pre-processing

E.A led the IHC and snRNAseq data generation

N.W led the IHC and snRNAseq data generation

M.P snRNAseq & IHC data generation

V.C IHC data generation

D.C IHC data generation

R.C.J M IHC data generation

H.W Proteomics data generation

R.Y Proteomics data generation

A.MG Bulk transcriptomics data generation

X.Z Bulk transcriptomics data generation

B.M.S Provided guidance on the analysis

J.J Supervised the study, conducted study design, analysis and interpretation of data

P.M Staff supervision, analysis and interpretation of data and funding All authors contributed, reviewed and approved the manuscript.

## References

1. Sheehan, B. Assessment scales in dementia. Ther Adv Neurol Disord 5, 349–358 (2012).

2. Morris, J. C. The Clinical Dementia Rating (CDR). Neurology 43, 2412.2-2412-a (1993).

3. Bennett, D. A. et al. Religious Orders Study and Rush Memory and Aging Project. Journal of Alzheimer’s Disease vol. 64 S161–S189 Preprint at 10.3233/JAD-179939 (2018).

4. Wang, M. et al. The Mount Sinai cohort of large-scale genomic, transcriptomic and proteomic data in Alzheimer’s disease. Sci Data 5, (2018).

5. La Cognata, V., Morello, G. & Cavallaro, S. Omics data and their integrative analysis to support stratified medicine in neurodegenerative diseases. International Journal of Molecular Sciences vol. 22 Preprint at 10.3390/ijms22094820 (2021).

6. Eteleeb, A. M. et al. Brain high-throughput multi-omics data reveal molecular heterogeneity in Alzheimer’s disease. PLoS Biol 22, e3002607 (2024).

7. De Strooper, B. & Karran, E. The Cellular Phase of Alzheimer’s Disease. Cell 164, 603–615 (2016).

8. Gerrits, E. et al. Distinct amyloid-β and tau-associated microglia profiles in Alzheimer’s disease. Acta Neuropathol (2021) doi:10.1007/s00401-021-02263-w.

9. Mancuso, R. et al. Xenografted human microglia display diverse transcriptomic states in response to Alzheimer’s disease-related amyloid-β pathology. Nat Neurosci (2024) doi:10.1038/s41593-024-01600-y.

10. Serrano-Pozo, A. et al. Title: Astrocyte transcriptomic changes along the spatiotemporal progression of Alzheimer’s disease Equal contribution. doi:10.1101/2022.12.03.518999.

11. Habib, N. et al. Disease-associated astrocytes in Alzheimer’s disease and aging. Nat Neurosci 23, 701–706 (2020).

12. Liddelow, S. A. et al. Neurotoxic reactive astrocytes are induced by activated microglia. Nature 541, 481–487 (2017).

13. Cameron, E. G. et al. A molecular switch for neuroprotective astrocyte reactivity. Nature 626, 574–582 (2024).

14. Sierksma, A. et al. Novel Alzheimer risk genes determine the microglia response to amyloid-β but not to TAU pathology. EMBO Mol Med 12, (2020).

15. Penney, J. et al. iPSC-derived microglia carrying the TREM2 R47H+ mutation are proinflammatory and promote synapse loss. Glia 72, 452–469 (2024).

16. Lawrence, J. M., Schardien, K., Wigdahl, B. & Nonnemacher, M. R. Roles of neuropathology-associated reactive astrocytes: a systematic review. Acta Neuropathol Commun 11, 42 (2023).

17. Liddelow, S. A. & Barres, B. A. Reactive Astrocytes: Production, Function, and Therapeutic Potential. Immunity 46, 957–967 (2017).

18. Matusova, Z., Hol, E. M., Pekny, M., Kubista, M. & Valihrach, L. Reactive astrogliosis in the era of single-cell transcriptomics. Front Cell Neurosci 17, (2023).

19. Livingston, N. R. et al. Relationship between astrocyte reactivity, using novel 11C-BU99008 PET, and glucose metabolism, grey matter volume and amyloid load in cognitively impaired individuals. Mol Psychiatry 27, 2019–2029 (2022).

20. Hulshof, L. A., van Nuijs, D., Hol, E. M. & Middeldorp, J. The Role of Astrocytes in Synapse Loss in Alzheimer’s Disease: A Systematic Review. Front Cell Neurosci 16, (2022).

21. Escartin, C. et al. Reactive astrocyte nomenclature, definitions, and future directions. Nat Neurosci 24, 312–325 (2021).

22. Paolicelli, R. et al. Defining Microglial States and Nomenclature: A Roadmap to 2030. SSRN Electronic Journal (2022) doi:10.2139/ssrn.4065080.

23. Argelaguet, R. et al. MOFA+: A statistical framework for comprehensive integration of multi-modal single-cell data. Genome Biol 21, (2020).

24. Skene, N. G. & Grant, S. G. N. Identification of vulnerable cell types in major brain disorders using single cell transcriptomes and expression weighted cell type enrichment. Front Neurosci 10, (2016).

25. Hodge, R. D. et al. Conserved cell types with divergent features in human versus mouse cortex. Nature 573, 61–68 (2019).

26. Fancy, N. et al. Mechanisms contributing to differential genetic risks for TREM2 R47H and R62H variants in Alzheimer’s Disease. doi:10.1101/2022.07.12.22277509.

27. Caramello, A. et al. Intra-cellular accumulation of amyloid is a marker of selective neuronal vulnerability in Alzheimer’s disease. doi:10.1101/2023.11.23.23298911.

28. de Leeuw, C. A., Mooij, J. M., Heskes, T. & Posthuma, D. MAGMA: Generalized Gene-Set Analysis of GWAS Data. PLoS Comput Biol 11, e1004219 (2015).

29. Bellenguez, C. et al. New insights into the genetic etiology of Alzheimer’s disease and related dementias. Nat Genet 54, 412–436 (2022).

30. Jansen, I. E. et al. Genome-wide meta-analysis identifies new loci and functional pathways influencing Alzheimer’s disease risk. Nat Genet 51, 404–413 (2019).

31. Wightman, D. P. et al. A genome-wide association study with 1,126,563 individuals identifies new risk loci for Alzheimer’s disease. Nat Genet 53, 1276– 1282 (2021).

32. Mo, Q. & Shen, R. IClusterPlus: Integrative Clustering of Multiple Genomic Data Sets. http://cran.r-project.org/web/packages/glmnet/index.html (2018).

33. Seyfried, N. T. et al. A Multi-network Approach Identifies Protein-Specific Co-expression in Asymptomatic and Symptomatic Alzheimer’s Disease. Cell Syst 4, 60–72.e4 (2017).

34. Honer, W. G. et al. Cognitive reserve, presynaptic proteins and dementia in the elderly. Transl Psychiatry 2, e114–e114 (2012).

35. Dammer, E. B. et al. Multi-platform proteomic analysis of Alzheimer’s disease cerebrospinal fluid and plasma reveals network biomarkers associated with proteostasis and the matrisome. Alzheimers Res Ther 14, 174 (2022).

36. Kodam, P., Sai Swaroop, R., Pradhan, S. S., Sivaramakrishnan, V. & Vadrevu, R. Integrated multi-omics analysis of Alzheimer’s disease shows molecular signatures associated with disease progression and potential therapeutic targets. Sci Rep 13, 3695 (2023).

37. Clark, C., Dayon, L., Masoodi, M., Bowman, G. L. & Popp, J. An integrative multi-omics approach reveals new central nervous system pathway alterations in Alzheimer’s disease. Alzheimers Res Ther 13, (2021).

38. Xicota, L. et al. Multi-omics signature of brain amyloid deposition in asymptomatic individuals at-risk for Alzheimer’s disease: The INSIGHT-preAD study. EBioMedicine 47, 518–528 (2019).

39. Peña-Bautista, C., Álvarez-Sánchez, L., Cañada-Martínez, A. J., Baquero, M. & Cháfer-Pericás, C. Epigenomics and lipidomics integration in alzheimer disease: Pathways involved in early stages. Biomedicines 9, (2021).

40. Klein, H. U., Schäfer, M., Bennett, D. A., Schwender, H. & de Jager, P. L. Bayesian integrative analysis of epigenomic and transcriptomic data identifies Alzheimer’s disease candidate genes and networks. PLoS Comput Biol 16, (2020).

41. Johnson, E. C. B. et al. Large-scale deep multi-layer analysis of Alzheimer’s disease brain reveals strong proteomic disease-related changes not observed at the RNA level. Nat Neurosci 25, 213–225 (2022).

42. Braak, H. & Braak, E. Acta H’ Pathologica Neuropathological Stageing of Alzheimer-Related Changes. Acta Neuropathol vol. 82 (1991).

43. Mathys, H. et al. Single-cell transcriptomic analysis of Alzheimer’s disease. Nature 570, 332–337 (2019).

44. Kumar, A. et al. Genetic effects on longitudinal cognitive decline during the early stages of Alzheimer’s disease. Sci Rep 11, 19853 (2021).

45. Yang, H. et al. Microglia-specific Alzheimer’s disease polygenic risk score is associated with amyloid-β, tau, and microglial activation. Alzheimer’s & Dementia 18, (2022).

46. Yang, H.-S. et al. Cell-type-specific Alzheimer’s disease polygenic risk scores are associated with distinct disease processes in Alzheimer’s disease. Nat Commun 14, 7659 (2023).

47. Lin, Y.-T. et al. APOE4 Causes Widespread Molecular and Cellular Alterations Associated with Alzheimer’s Disease Phenotypes in Human iPSC-Derived Brain Cell Types. Neuron 98, 1141–1154.e7 (2018).

48. Therriault, J. et al. Biomarker modeling of Alzheimer’s disease using PET-based Braak staging. Nat Aging 2, 526–535 (2022).

49. Wang, X., Ye, T., Zhou, W. & Zhang, J. Uncovering heterogeneous cognitive trajectories in mild cognitive impairment: a data-driven approach. Alzheimers Res Ther 15, 57 (2023).

50. Leng, K. et al. Molecular characterization of selectively vulnerable neurons in Alzheimer’s disease. Nat Neurosci 24, 276–287 (2021).

51. DeKosky, S. T. & Scheff, S. W. Synapse loss in frontal cortex biopsies in Alzheimer’s disease: Correlation with cognitive severity. Ann Neurol 27, 457–464 (1990).

52. Williams, J. B., Cao, Q. & Yan, Z. Transcriptomic analysis of human brains with Alzheimer’s disease reveals the altered expression of synaptic genes linked to cognitive deficits. Brain Commun 3, (2021).

53. Zhang, H. et al. Cerebrospinal fluid synaptosomal-associated protein 25 is a key player in synaptic degeneration in mild cognitive impairment and Alzheimer’s disease. Alzheimers Res Ther 10, 80 (2018).

54. Giuffrè, G. M. et al. Cerebrospinal fluid neurofilament light chain and total-tau as biomarkers of neurodegeneration in Alzheimer’s disease and frontotemporal dementia. Neurobiol Dis 186, 106267 (2023).

55. Qiang, Q., Skudder-Hill, L., Toyota, T., Wei, W. & Adachi, H. CSF GAP-43 as a biomarker of synaptic dysfunction is associated with tau pathology in Alzheimer’s disease. Sci Rep 12, 17392 (2022).

56. Hakobyan, S. et al. Complement Biomarkers as Predictors of Disease Progression in Alzheimer’s Disease. Journal of Alzheimer’s Disease 54, 707–716 (2016).

57. Plassman, B. L. et al. Incidence of dementia and cognitive impairment, not dementia in the united states. Ann Neurol 70, 418–426 (2011).

58. Valdés, A. et al. Proteomic comparison between different tissue preservation methods for identification of promising biomarkers of urothelial bladder cancer. Sci Rep 11, 7595 (2021).

59. Kong, A. T., Leprevost, F. V, Avtonomov, D. M., Mellacheruvu, D. & Nesvizhskii, A. I. MSFragger: ultrafast and comprehensive peptide identification in mass spectrometry–based proteomics. Nat Methods 14, 513–520 (2017).

60. Demichev, V., Messner, C. B., Vernardis, S. I., Lilley, K. S. & Ralser, M. DIA-NN: neural networks and interference correction enable deep proteome coverage in high throughput. Nat Methods 17, 41–44 (2020).

61. Smith, A. M. et al. Diverse human astrocyte and microglial transcriptional responses to Alzheimer’s pathology. Acta Neuropathol 143, 75–91 (2022).

62. Krishnaswami, S. R. et al. Using single nuclei for RNA-seq to capture the transcriptome of postmortem neurons. Nat Protoc 11, 499–524 (2016).

63. Fancy, N. N. et al. Characterisation of premature cell senescence in Alzheimer’s disease using single nuclear transcriptomics. Acta Neuropathol 147, 78 (2024).

64. Schneegans, E. et al. Omix: A Transcriptomics-Proteomics Integration Pipeline. (2023).

65. Van den Berge, K., et al. Trajectory-based differential expression analysis for single-cell sequencing data. Nat Commun 11, 1201 (2020).

66. Street, K. et al. Slingshot: cell lineage and pseudotime inference for single-cell transcriptomics. BMC Genomics 19, 477 (2018).

67. Blondel, V. D., Guillaume, J.-L., Lambiotte, R. & Lefebvre, E. Fast unfolding of communities in large networks. Journal of Statistical Mechanics: Theory and Experiment 2008, P10008 (2008).

68. Ritchie, M. E. et al. limma powers differential expression analyses for RNA-sequencing and microarray studies. Nucleic Acids Res 43, e47–e47 (2015).

69. Benjamini, Y. & Hochberg, Y. Controlling the False Discovery Rate: A Practical and Powerful Approach to Multiple Testing. J R Stat Soc Series B Stat Methodol 57, 289–300 (1995).

70. Chen, E. Y. et al. Enrichr: interactive and collaborative HTML5 gene list enrichment analysis tool. BMC Bioinformatics 14, 128 (2013).

71. Morabito, S., Reese, F., Rahimzadeh, N., Miyoshi, E. & Swarup, V. hdWGCNA identifies co-expression networks in high-dimensional transcriptomics data. Cell Reports Methods 3, 100498 (2023).

72. Keenan, A. B. et al. ChEA3: transcription factor enrichment analysis by orthogonal omics integration. Nucleic Acids Res 47, W212–W224 (2019).

73. Purcell, S. et al. PLINK: A Tool Set for Whole-Genome Association and Population-Based Linkage Analyses. The American Journal of Human Genetics 81, 559–575 (2007).

74. Chalise, P. & Fridley, B. L. Integrative clustering of multi-level ‘omic data based on non-negative matrix factorization algorithm. PLoS One 12, e0176278 (2017).

75. Meng, C., Helm, D., Frejno, M. & Kuster, B. moCluster: Identifying Joint Patterns Across Multiple Omics Data Sets. J Proteome Res 15, 755–765 (2016).

76. Bates, D., Mächler, M., Bolker, B. & Walker, S. Fitting Linear Mixed-Effects Models Using **lme4**. J Stat Softw 67, (2015).

