## Supplementary figures for "Integrative multi-omics reveal glial signatures associated with accelerated cognitive decline in Alzheimer’s disease"

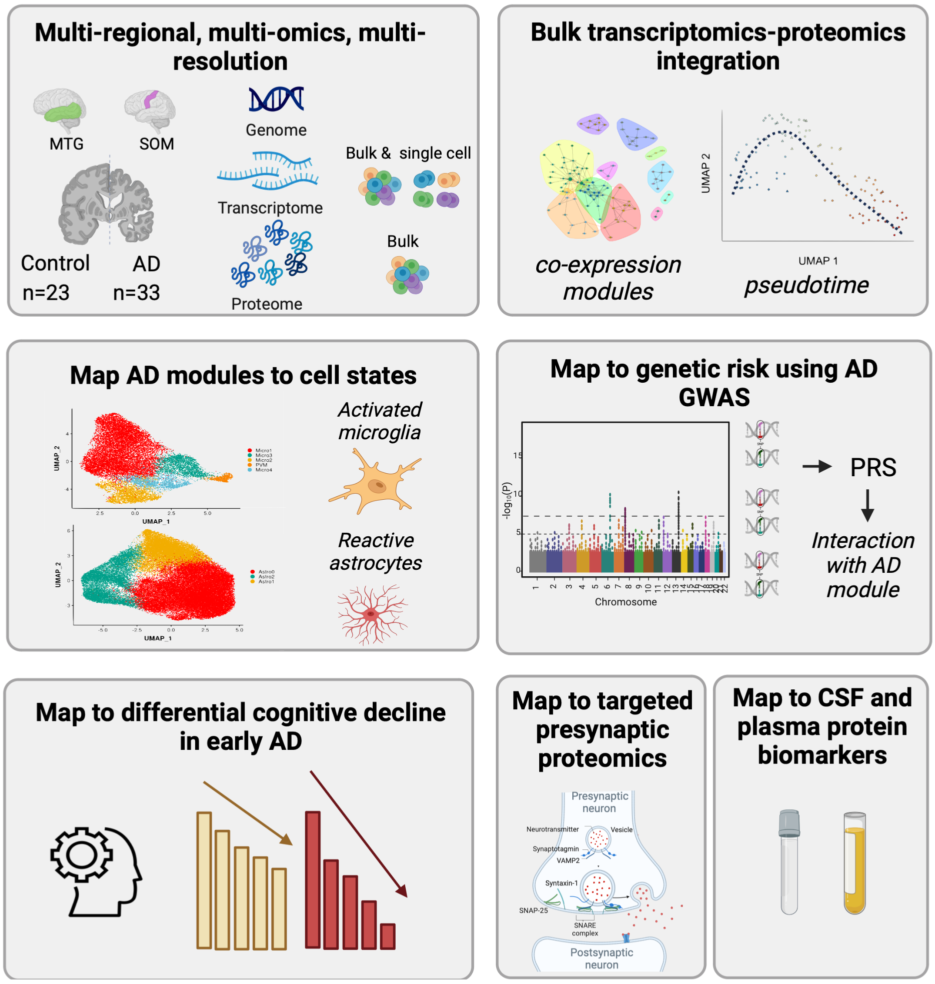

S1 Fig. A Comprehensive Multi-Regional, Multi-Omic, Multi-Resolution analysis

**a)** Multi-regional analysis depicting samples from control and AD patients from the middle temporal gyrus (MTG) and somatosensory cortex (SOM), integrating genomic, transcriptomic (both bulk and single-cell), and bulk proteomics data. **b)** Integration of bulk RNA and proteomics profiling, generation of co-expression modules and derivation of pseudo-time. **c)** Projecting bulk modules on single-cell transcriptomics to identify cell state-specific AD modules emphasising activated microglia and reactive astrocytes. **d)** Genetic susceptibility to AD was mapped using polygenic risk scores (PRS) derived from GWAS and their interaction with identified AD modules. **e)** Correlation of these modules with differential cognitive impairment. **f)** Targeted SNARE proteomics reflecting synaptic functionality. **g)** Correlation of AD-related modules with CSF and plasma protein biomarkers.

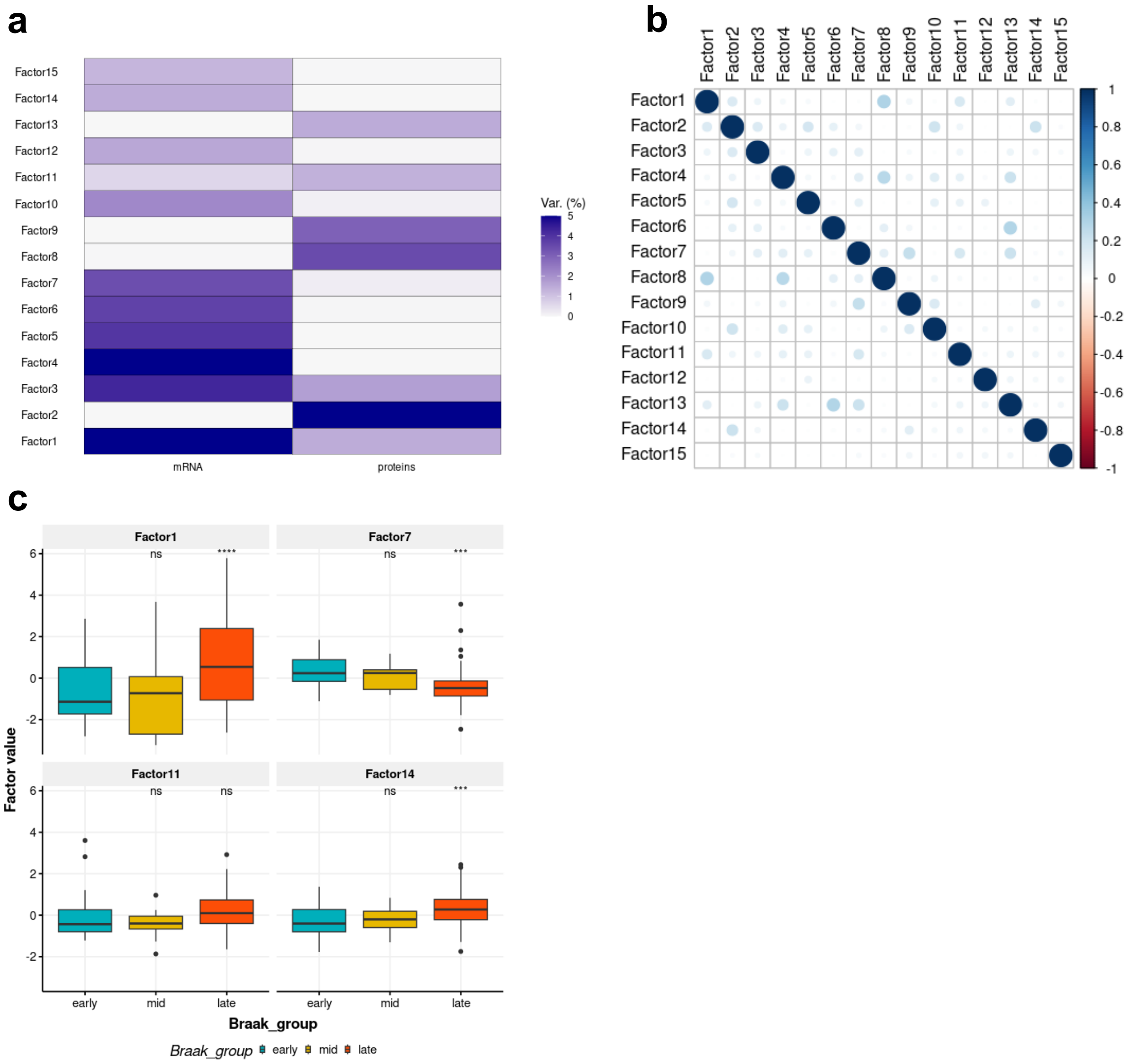

S2 Fig. Transcriptomics-proteomics integration using MOFA

**a)** Variance explained by the learnt factors across transcriptomics and proteomics**. b)** Factors correlation matrix where poor correlation between factors suggests a good model fit. **c)** AD-relevant factors differences between Mid (Braak III-IV) and late (Braak V-VI) relative to early (Braak 0-II) (*Two-sided* Wilcoxon rank-sum test, adjusted p, *p<=0.05, **p<=0.01, ***p<=0.001)

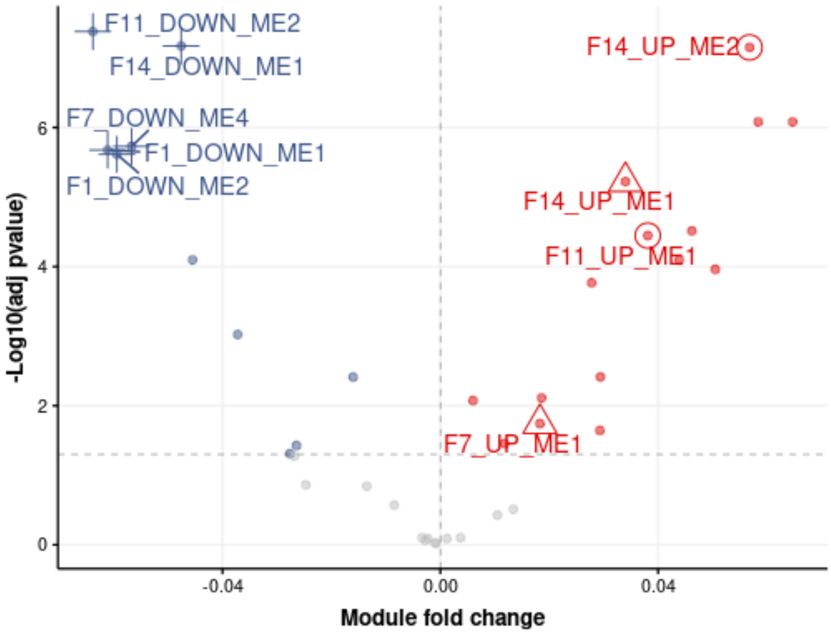

S3 Fig. Module eigenvalue differential expression against pseudotime

**a)** Volcano plot illustrating module fold changes plotted against pseudotime, accounting for various confounding factors, with up-regulated (red) and down-regulated (blue) significant modules labelled. Circles, triangles, and crosses indicate astrocyte, microglial and neuronal enrichment, respectively.

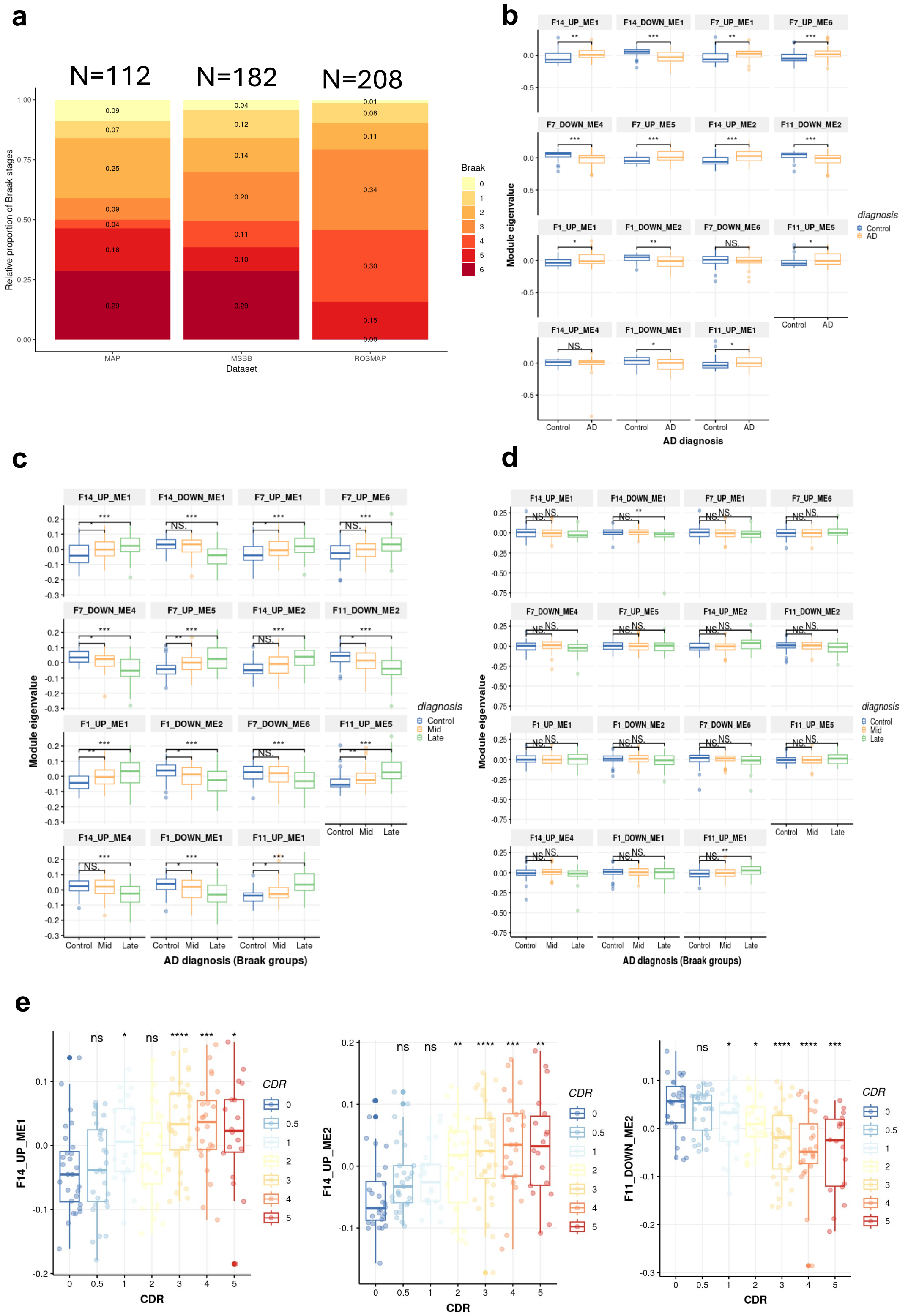

S4 Fig. Module eigenvalue validation in external cohorts (ROSMAP, MSBB)

**a)** Discovery cohort (MAP) and validation cohorts (MSBB, ROSMAP) repartition of samples by Braak stages. MSBB and MAP show a similar Braak distribution, while ROSMAP has a higher representation of Mid Braak (III-IV). **b)** Module differences in the discovery cohort. **c)** Module differences in MSBB, demonstrating consistent patterns of up-regulation and down-regulation of module eigenvalues in AD compared to controls, similar to those observed in the discovery cohort. **d)** Module differences in ROSMAP do not show significant differences, which is attributed to the cohort composition (more middle stage and fewer early and late-stage cases). **e)** CDR differences in microglia, astrocyte and neuronal injury modules in MSBB. (*Two-sided* Wilcoxon rank-sum test, adjusted p, *p<=0.05, **p<=0.01, ***p<=0.001)

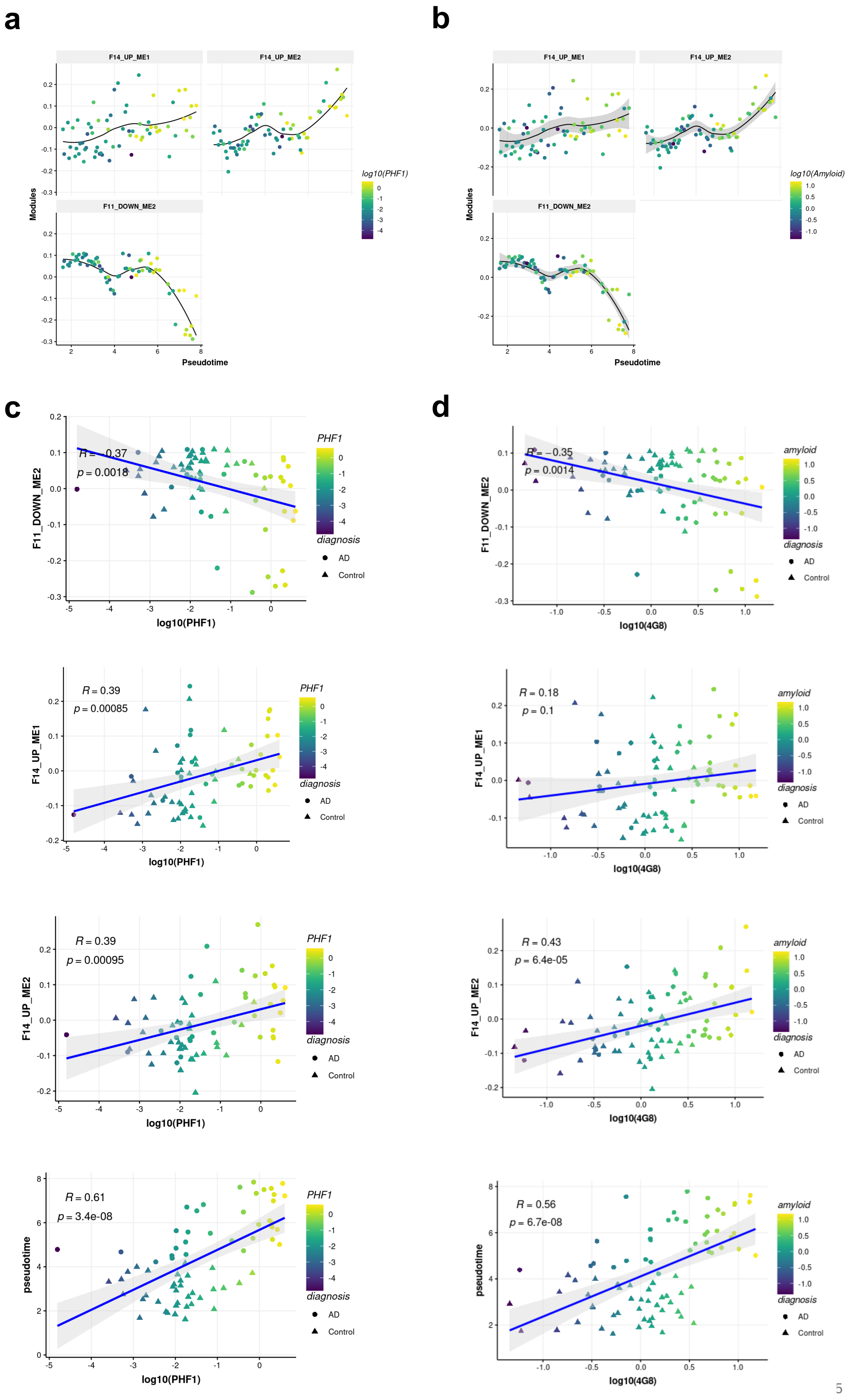

S5 Fig. Module trajectories alongside pseudotime and association to neuropathology

**a)** Module eigenvalue as a function of pseudotime (loess regression). Points coloured by log10(PHF1) **b)** Same as a) but points are colours by log10(amyloid) load. **c)** Synaptic module, microglia, astrocyte module eigenvalue and pseudotime as a function of log10(PHF1) **d)** Same as in c) but as a function of log10(amyloid) measured by 4G8.

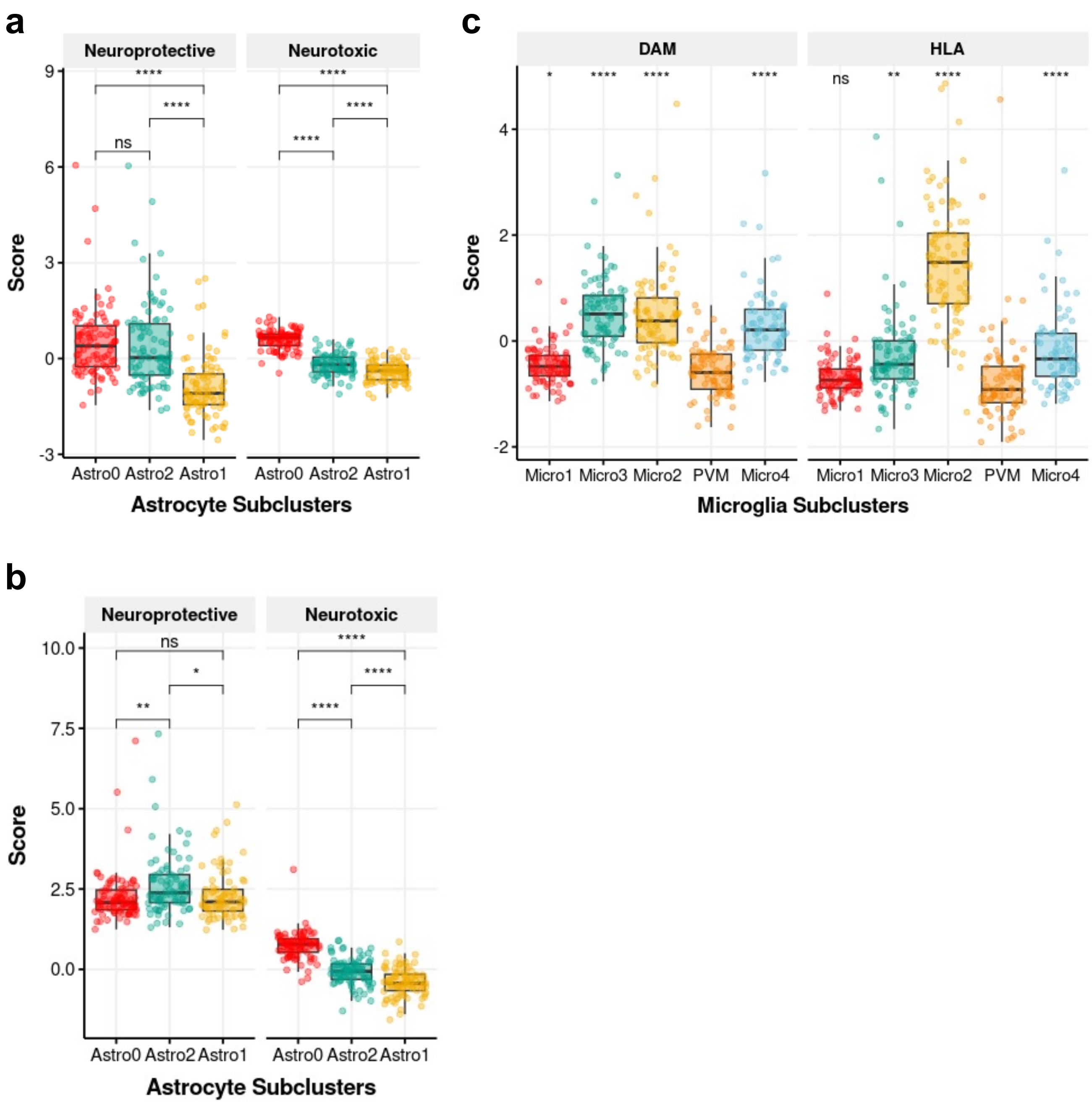

S6 Fig. Sample level gene set scores in astrocyte and microglia subpopulations

**(a)** Scores for neuroprotective and neurotoxic states across astrocyte subclusters (Astro0, Astro2, Astro1). The neuroprotective scores show significant differences with Astro2 and Astro1 having significantly higher scores compared to Astro0. For neurotoxic scores, Astro0 shows a significantly higher profile **(b)** Similar to panel (a), this panel shows results after applying 75$^{th}$ percentile thresholding on neuroprotective scores. Neurotoxic scores are filtered to values that are higher to the neuroprotective score. The results are consistent with panel (a), highlighting that only Astro2 is enriched in neuroprotective genes when considering the top 25% neuroprotective values. **(c)** Scores for DAM and HLA across six microglia subclusters (Micro1, Micro3, Micro2, PVM, Micro4). Pairwise tests are performed relative to the PVM cluster. (*t.test*, **p<=0.05, **p<=0.01, ***p<=0.001*)

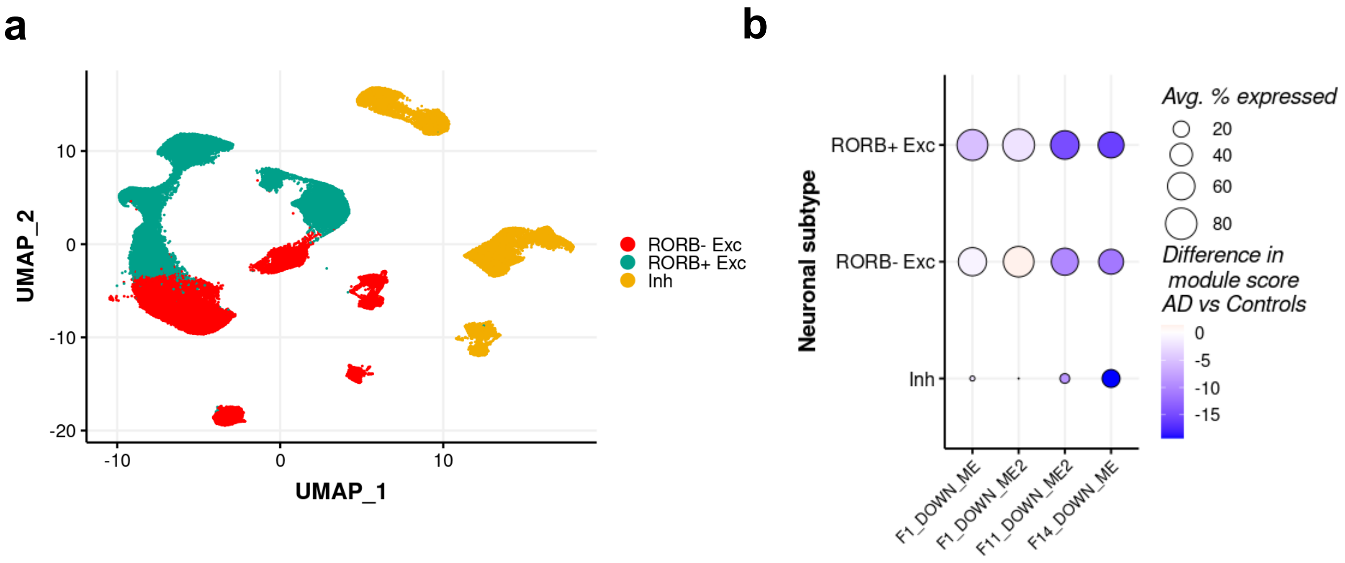

S7 Fig. Neuronal subpopulations and neuronal bulk module projections

**a)** UMAP visualisation of neuronal subtypes in snRNAseq data grouped by RORB positivity in excitatory subtypes. **b)** Projecting AD-related neuronal modules on single-cell clusters using HdWGCNA grouped by RORB positivity in excitatory subtypes.

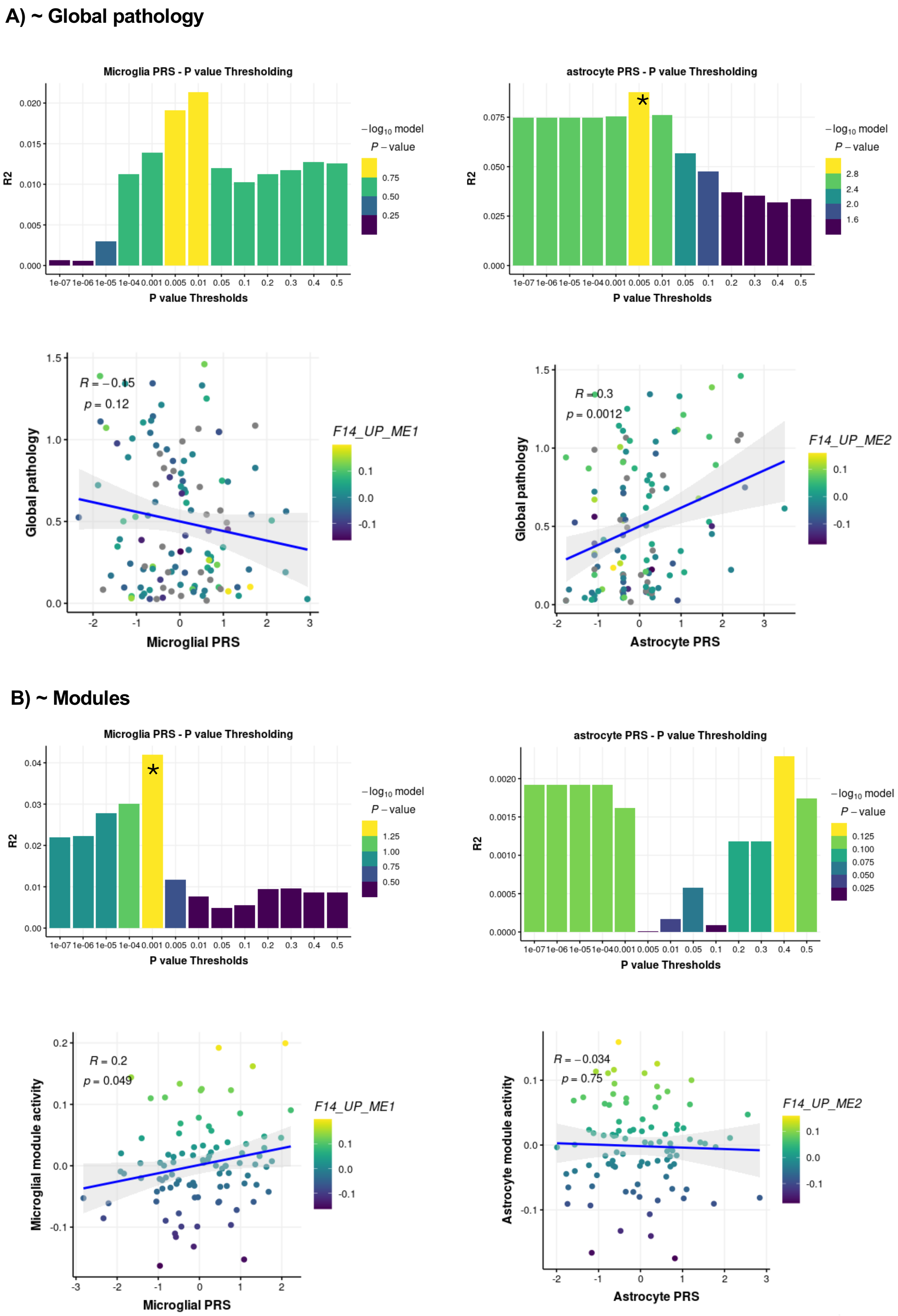

S8 Fig. P-value thresholding for microglia and astrocyte PRS

**a)** P-value thresholding according to global pathology. Astrocyte PRS R2 was maximised when modelled against global pathology, with a best-fit PRS at the 0.005 threshold (denoted by *). **b)** P-value thresholding according to microglia/astrocyte module eigenvalues. Microglia PRS R2 was maximised when modelled against microglia module activity with a best-fit PRS at the 0.001 threshold (denoted by *).

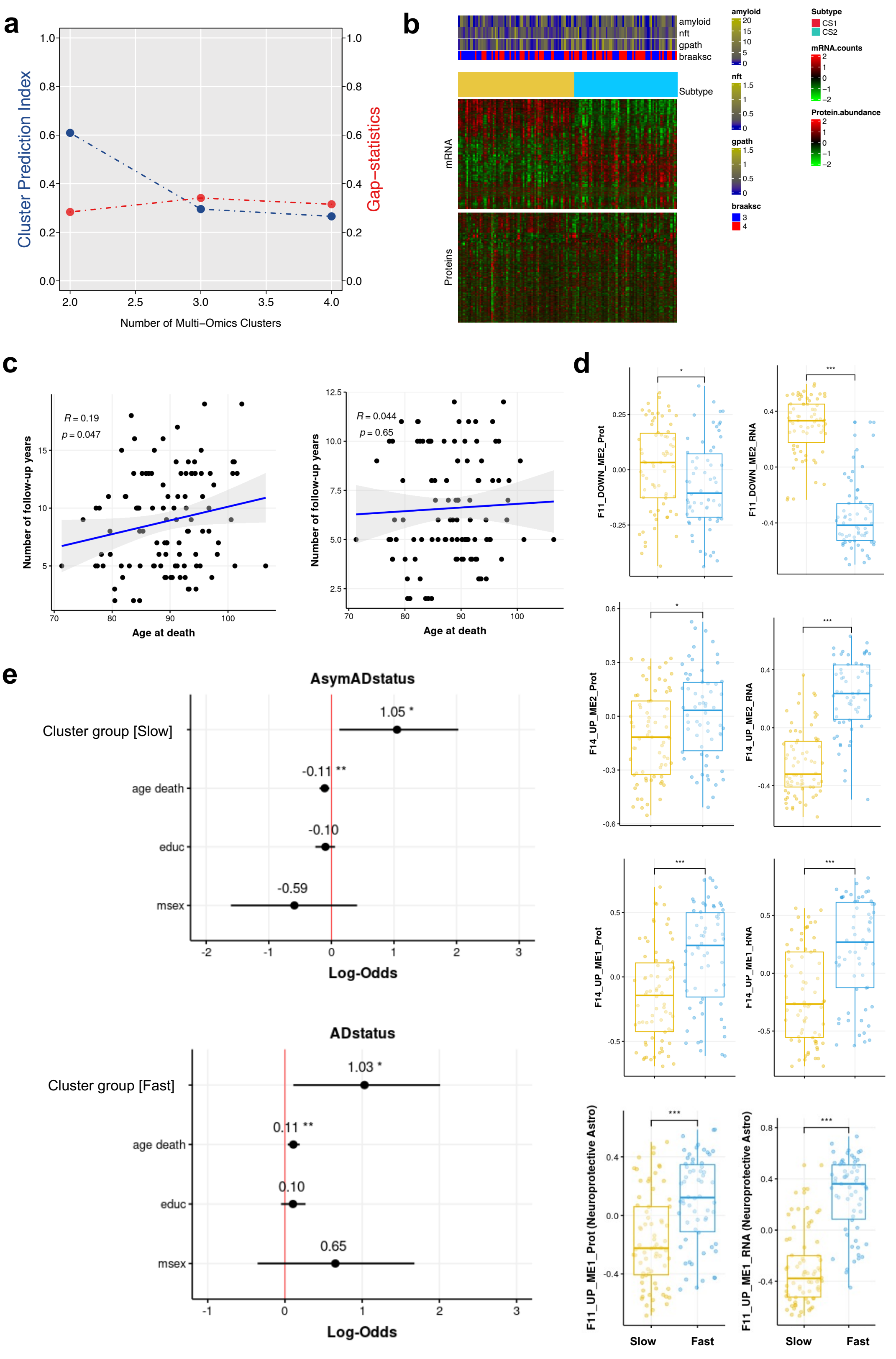

#### S9 Fig. ROSMAP cohort in early AD – multi-omics clustering

**a)** Cluster number optimisation: The optimal number of clusters was determined by combining the CPI and Gap-statistics results, selecting the number that maximised the sum of the CPI score and Gap statistic (k=2). **b)** Multi-omics clusters transcriptomics and proteomics expression heatmap. **c)** Relationship between number of follow-up years as a function of age at death to detect survivor bias in early AD ROSMAP cohort (n=132). Bias is corrected for when filtering for participants with less than 12 years of follow-up (n=102). **d)** Module eigenvalue differences in different modules are separated by proteomics and transcriptomics. (*Two-sided* Wilcoxon rank-sum test, adjusted p, *p<=0.05, **p<=0.01, ***p<=0.001) **e)** Logistic regression analysis assessing the influence of cluster group on AD symptomatic (MMSE<25) or asymptomatic (MMSE>=25) status, adjusted for age at death, education level, and sex.

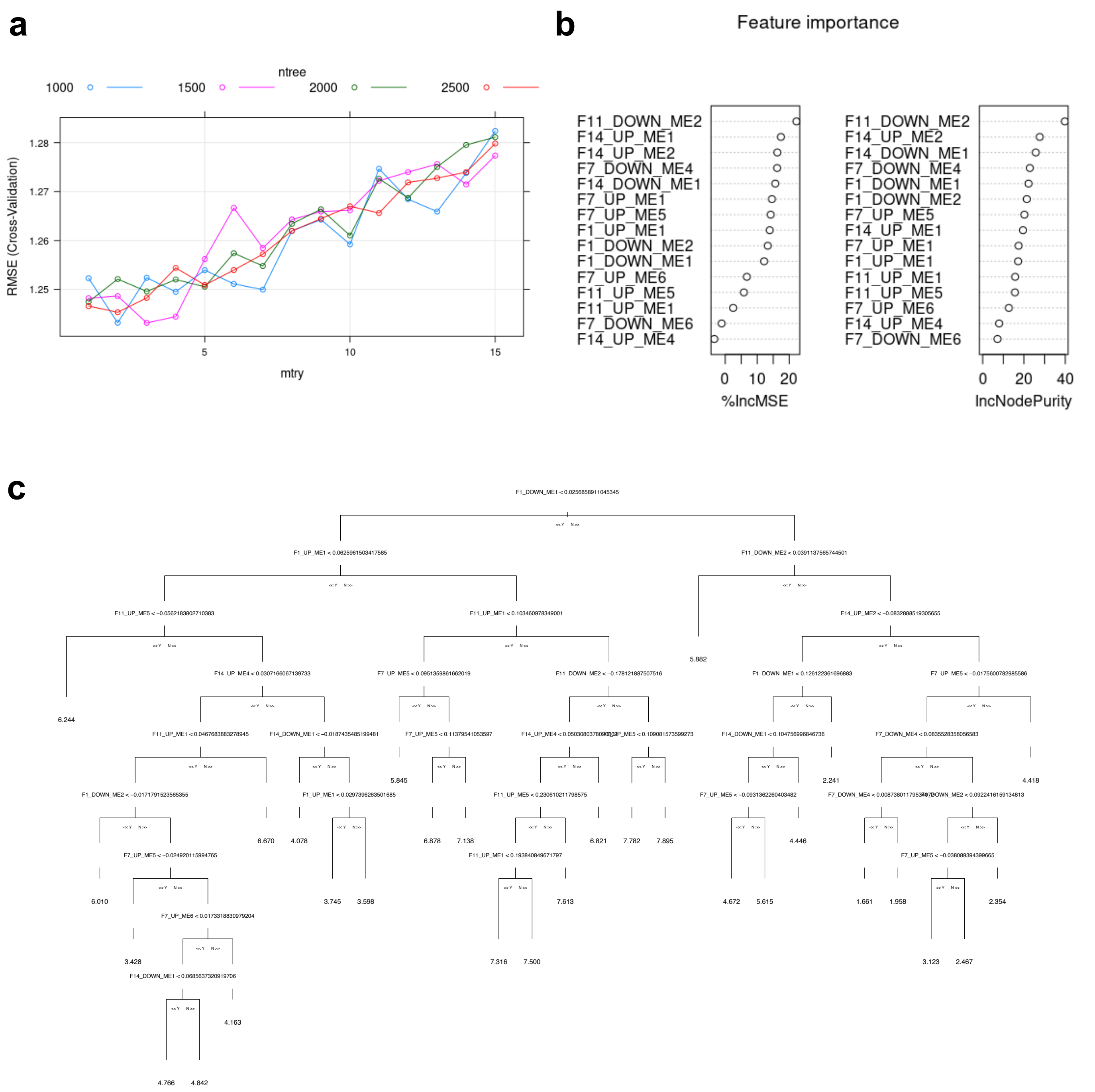

### S10 Fig. Random forest model to predict pseudotime

**a)** Hyperparameter tuning results for the random forest model. The graph shows the RMSE values obtained through 10-fold cross-validation, for different values of mtry (number of variables randomly sampled as candidates at each split) and ntree (number of trees in the forest). The coloured lines represent different ntree values (1000, 1500, 2000, 2500). The optimal hyperparameters were identified as mtry = 3 and ntree = 1500, indicated by the lowest RMSE values. **b)** Variable importance plots for the random forest model. The left plot shows the percent increase in mean squared error (%IncMSE) when each variable is permuted, indicating the impact of each variable on model accuracy. The right plot shows the increase in node purity (IncNodePurity) for each variable, representing their contribution to reducing impurity in the decision trees. Both plots highlight the most influential features used in predicting pseudotime. **c)** Tree illustrating the decision rules and splits made based on different variables and their thresholds. The tree nodes display the variable names, split points, and the predicted pseudotime values at the terminal nodes (leaves).
