## Supplementary material for "Integrative multi-omics reveal glial signatures associated with accelerated cognitive decline in Alzheimer’s disease": Reports: proteomics_vulnerable_DOWN.html

Omix - Pathway report


### Pathway Report


### **Omix** - Pathway report

###### 14 May, 2024

#### Pathway report overview

- **Functional enrichment**
- **Enrichment database:** GO\_Molecular\_Function\_2021, GO\_Cellular\_Component\_2021, GO\_Biological\_Process\_2021, Reactome\_2016, KEGG\_2021\_Human, MSigDB\_Hallmark\_2020, BioCarta\_2016

### Functional enrichment results

#### Data tables

$GO\_Molecular\_Function\_2021

$GO\_Cellular\_Component\_2021

$GO\_Biological\_Process\_2021

$Reactome\_2016

$KEGG\_2021\_Human

$MSigDB\_Hallmark\_2020

$

### Functional enrichment plot

#### Pathways

$GO\_Molecular\_Function\_2021  $GO\_Cellular\_Component\_2021  $GO\_Biological\_Process\_2021  $Reactome\_2016  $KEGG\_2021\_Human  $MSigDB\_Hallmark\_2020  $ NULL

### Functional enrichment results as networks

#### Networks

Omix v1.0.0 – 2024-05-14 12:40:59

---

A report by **Omix**
