## Supplementary material for "Integrative multi-omics reveal glial signatures associated with accelerated cognitive decline in Alzheimer’s disease": Reports: F11_DOWN_2.html

Omix - Pathway report


### Quality control report


### **Omix** - Pathway report

###### 06 December, 2023

#### Pathway report overview

- **Functional enrichment**
- **Enrichment database:** GO\_Molecular\_Function\_2021,
  GO\_Cellular\_Component\_2021, GO\_Biological\_Process\_2021, Reactome\_2016,
  KEGG\_2021\_Human, MSigDB\_Hallmark\_2020

### Functional enrichment results

#### Data tables

$GO\_Molecular\_Function\_2021

$GO\_Cellular\_Component\_2021

$GO\_Biological\_Process\_2021

$Reactome\_2016

$KEGG\_2021\_Human

$MSigDB\_Hallmark\_2020

### Functional enrichment plot

#### Pathways

$GO\_Molecular\_Function\_2021
$GO\_Cellular\_Component\_2021
$GO\_Biological\_Process\_2021
$Reactome\_2016
$KEGG\_2021\_Human
$MSigDB\_Hallmark\_2020

### Functional enrichment results as networks

#### Networks

Omix v1.0.0 – 2023-12-06 15:00:02

---

A report by **Omix**
